## Supplementary Appendix for "How geographic access to care shapes disease burden: the current impact of post-exposure prophylaxis and potential for expanded access to prevent human rabies deaths in Madagascar"

Malavika Rajeev et al.

### S1. Estimating travel times to the closest clinic provisioning PEP

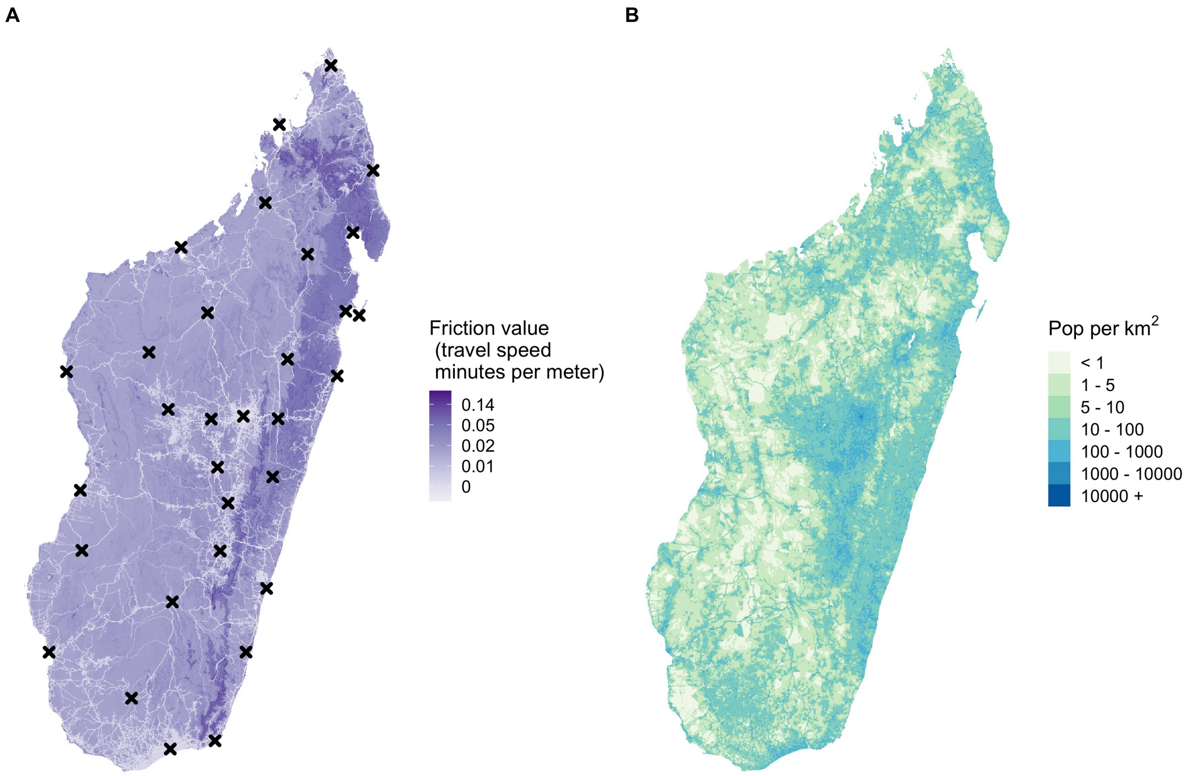

***Fig S1.1. Raster inputs to estimate travel times to the closest clinic provisioning PEP for Madagascar.***

*(A) Friction surface of travel speeds (in minutes per meter) at an ~ 1x1 km scale, with location of current clinics provisioning PEP (N = 31) shown with black x's. (B) Population estimates resampled to the same friction surface.*

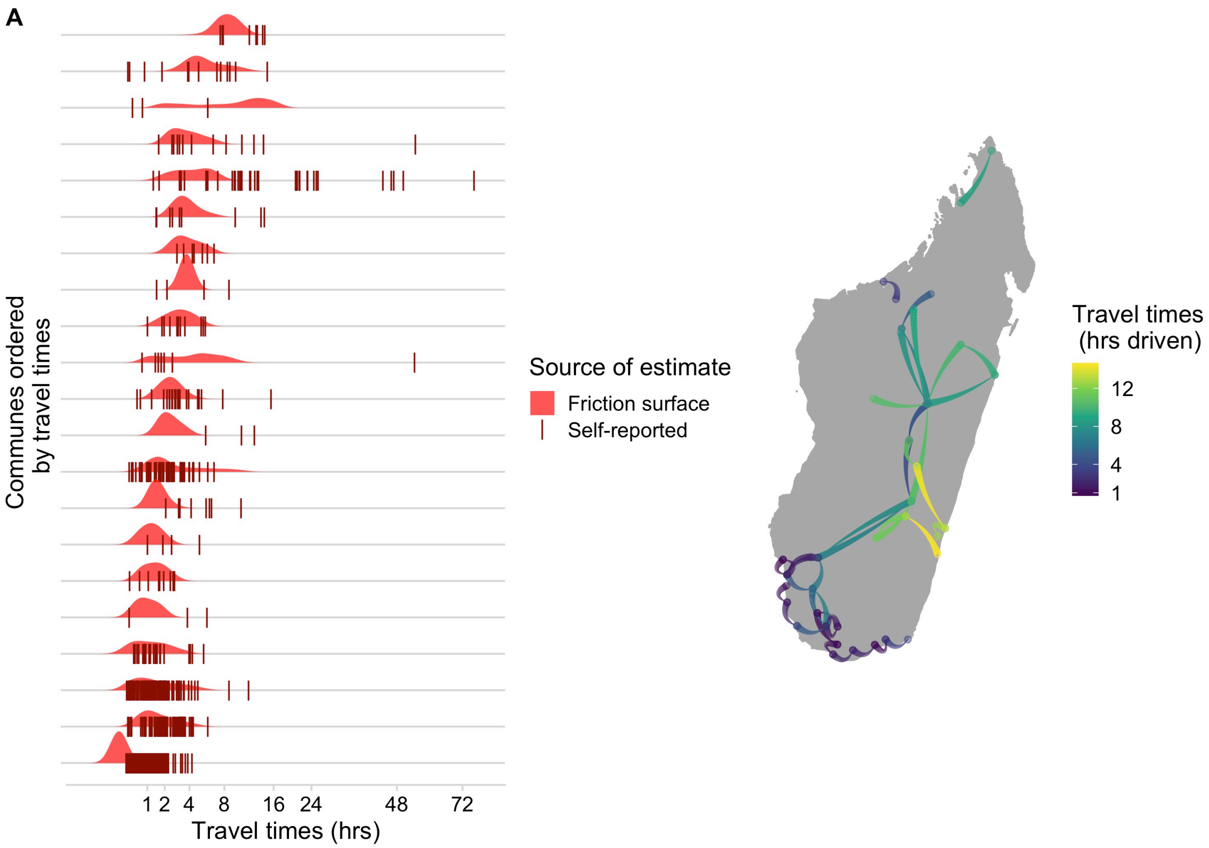

***Fig S1.2. Raw patient reported and driving travel time data.***

*(A) Distribution of travel times estimated at the grid cell level and reported by patients for each commune where patient data were available from the Moramanga PEP clinic (B) Reported driving times between locations, with the color corresponding to the total driving time and the size of the line showing the direction of travel (narrow to wide ~ origin to destination). Paths are Bezier curves from origin to destination, and do not show actual paths driven.*

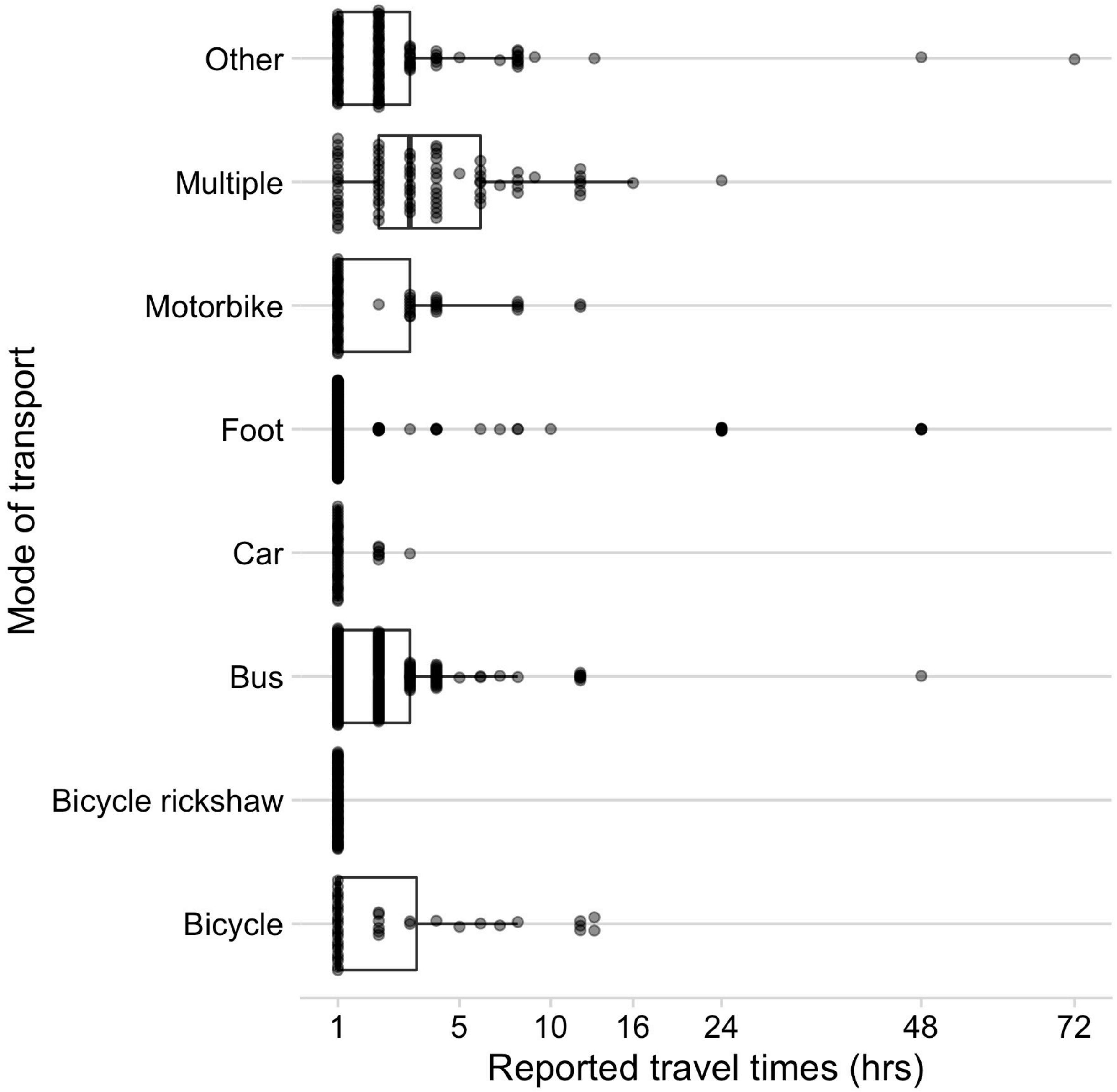

***Fig S1.3. Reported modes of transport used compared to reported travel times for patients reporting to the Moramanga PEP clinic***

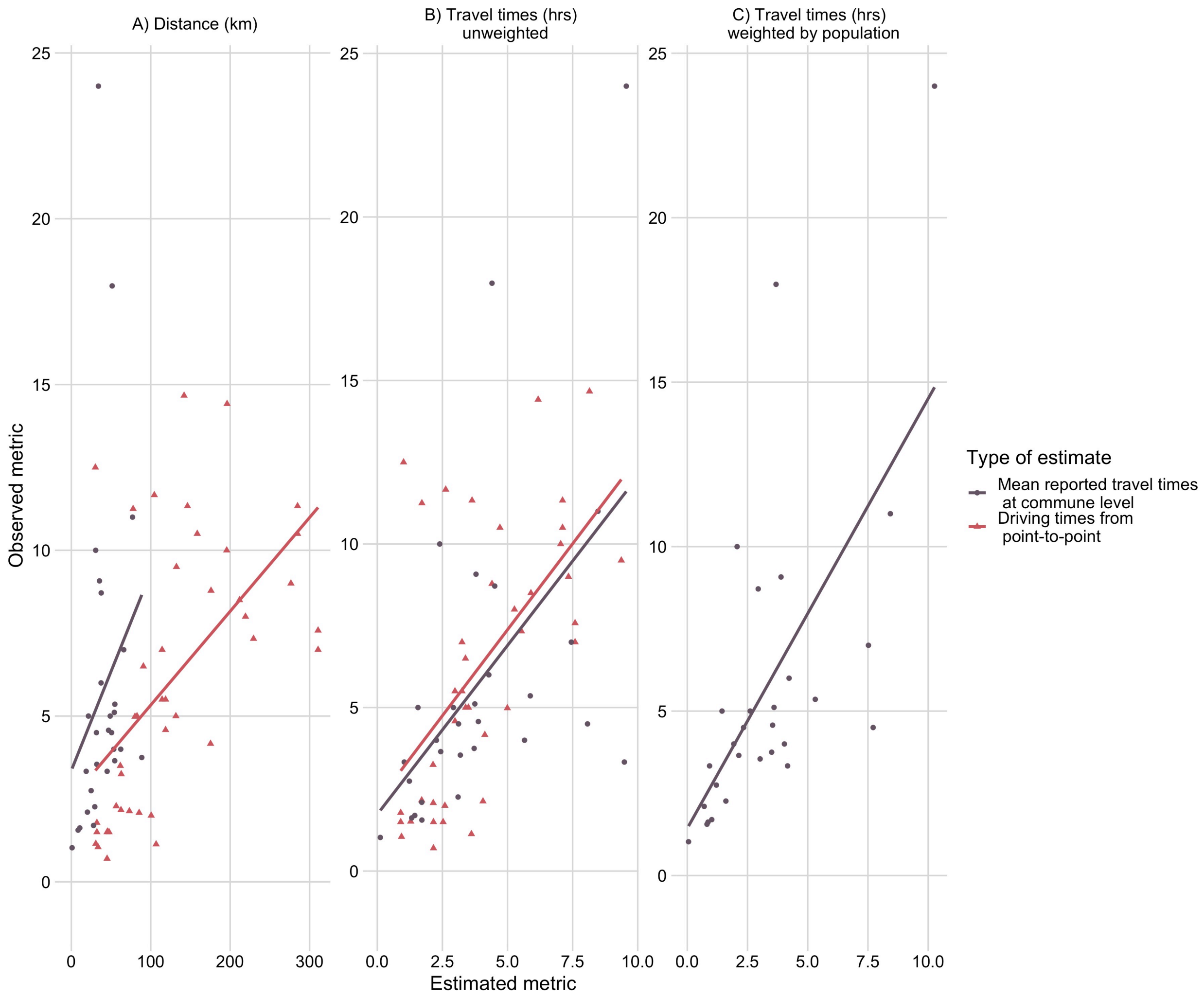

***Fig S1.4. Observed estimates of travel times (commune means of patient reported travel times and driving times between point locations) vs. estimates from friction surface.***

*Predicted by (A) Distance (km) (Euclidean distance between origin and destination for driving times and distance from the commune centroid to the Moramanga PEP clinic for commune means) (B) Travel time estimates and (C) Travel time estimates weighted by population (for commune means only) .*

***Table S1.1. $R^2$ values from linear models with estimated access metrics predicting either driving times or commune means of patient reported travel times.***

| Predictor | Driving times | Commune mean of patient reported travel times |
|---|---|---|
| Weighted travel times (hrs) | NA | 0.433 |
| Unweighted travel times (hrs) | 0.347 | 0.290 |
| Distance (km) | 0.368 | 0.093 |

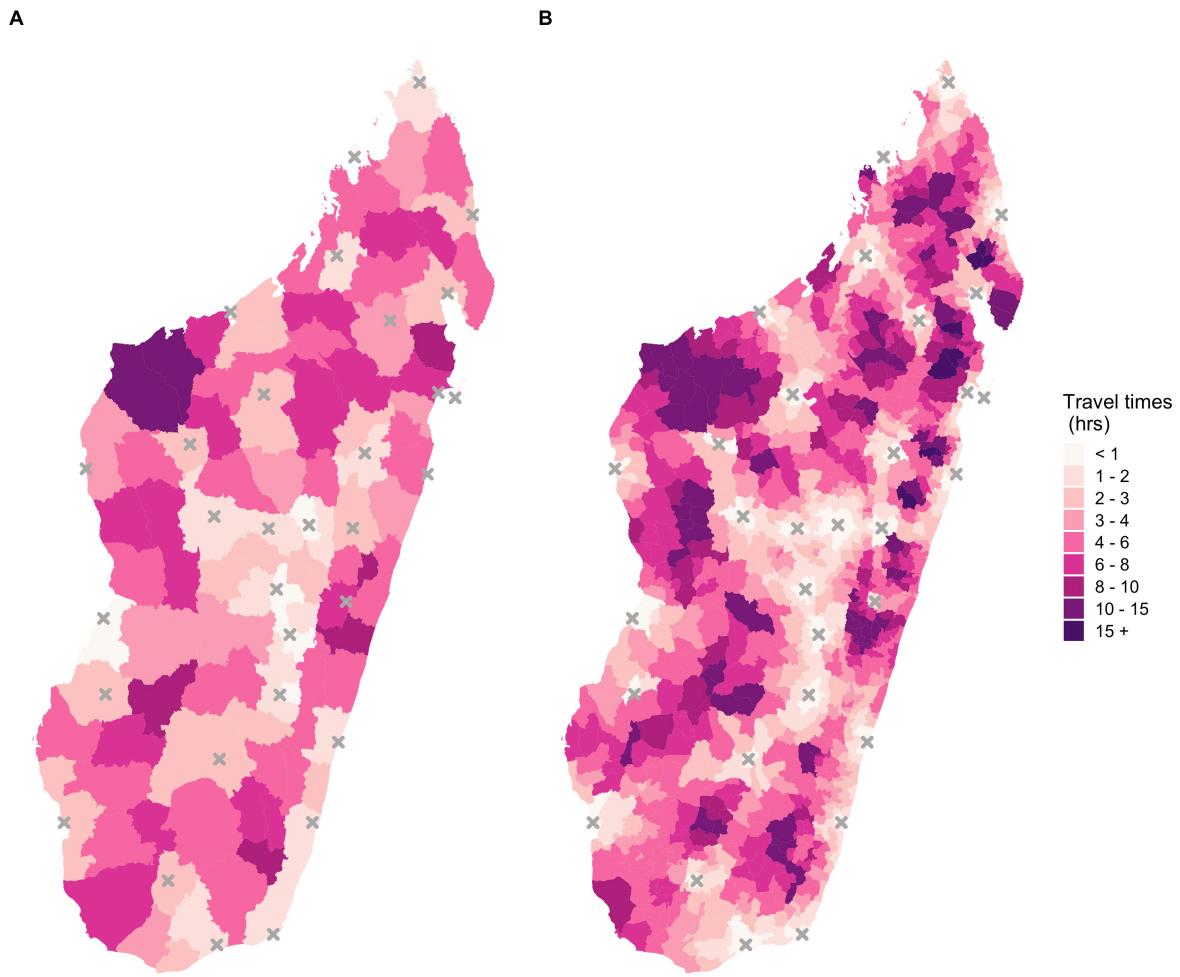

***Fig S1.5. Estimates of mean travel times weighted by population at the (A) District (B) Commune scale.***

**S2. Estimating bite incidence**

Most patients from each district reported to their closest clinic provisioning PEP by the weighted travel time metric (Fig 3). Accordingly, we assigned catchments based on which clinic was the closest for the majority of the population. While there are discrepancies between commune and district catchment assignments (Fig S2.1A), over 75% of the population in a given district or commune were closest to a single clinic (Fig S2.1C). We excluded any clinics which submitted less than 10 forms (excluded 11 catchments, Fig 3A grey polygons) and corrected for periods where clinics did not submit any forms.

Vial demand was simulated under simplified assumptions of PEP administration and adherence [1], based on patients reported randomly across the year. During this period, the Thai Red Cross Intradermal regimen was used across Madagascar, with 0.2 mL administered per patient completing doses on days 0, 3, 7, and 28. Vials can be shared within a day between two patients, resulting in 0.1ml wastage per vial shared, plus any additional wastage from unused doses discarded at the end of the day. We estimate vial estimates as the midpoint estimate if all patients complete 3 vs. 4 doses. As clinic submission of forms was highly variable from 2014-2017 (Fig S2.2A), we compared estimated demand to the total vials provisioned across this four year period comparing different thresholds for correcting for periods of no form submission (i.e, designating periods of 1, 5, 10, 15, and 30 consecutive days with zero submitted records as missing, compared to no correction).

Estimates of vial demand based on uncorrected bite patient numbers were generally lower than the number of vials provisioned for those clinics with substantial under-submission of forms (Fig S2.2B). Correcting patient numbers for under-submission resulted in estimates of vial demand closer to the provisioned vials for most clinics (Fig S2.2C, Table S2.1).

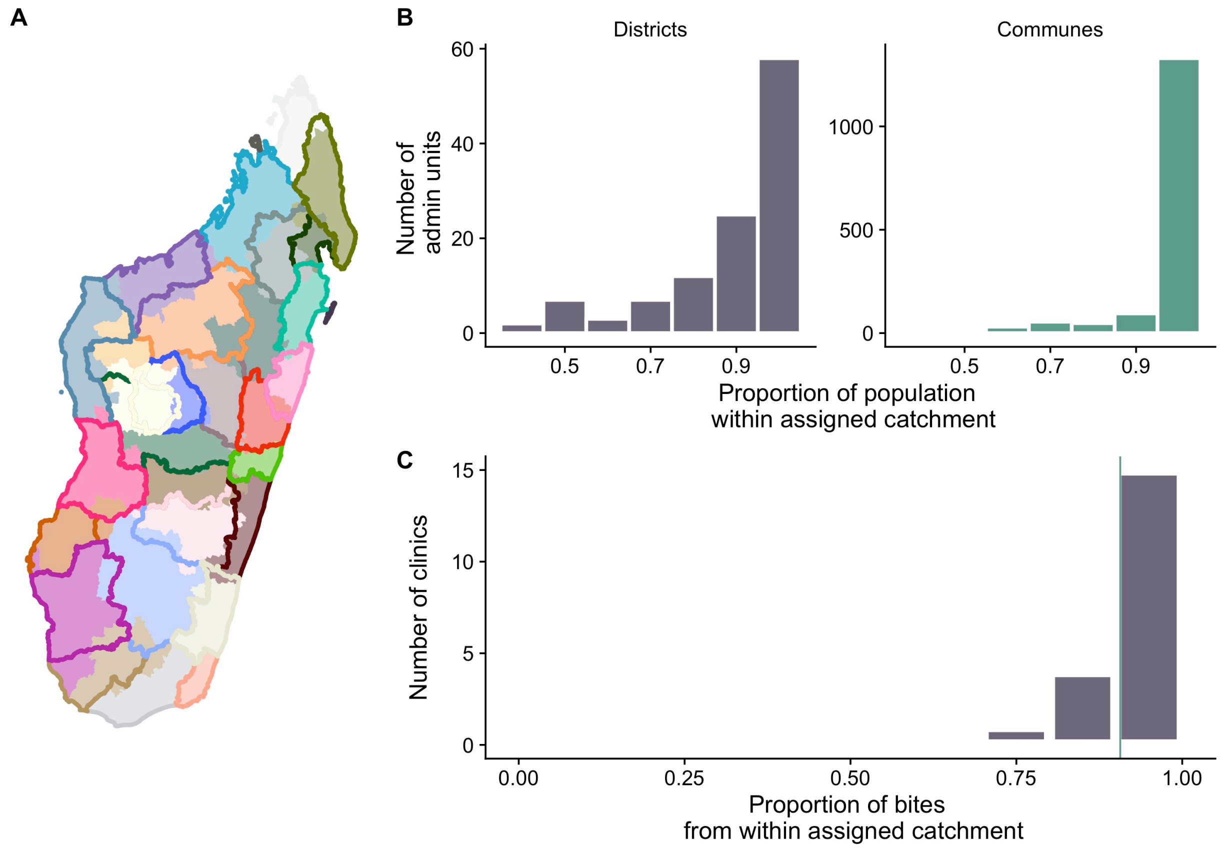

***Fig S2.1. Catchment assignments by travel time.***

*(A) Catchments as assigned by closest clinic for the majority of the population within a commune (polygon fill) or within a district (polygon outline). Admin units where the fill and border colors do not match show places where assigned catchments differ at the district vs commune scale. (B) Distribution of the proportion of the population in a given administrative unit (district or commune) served by the catchment assigned. (C) The proportion of bites reported to each clinic which originated from a district within the assigned catchment. The vertical line indicates the proportion of bites from within the assigned catchment for the Moramanga data (~90%).*

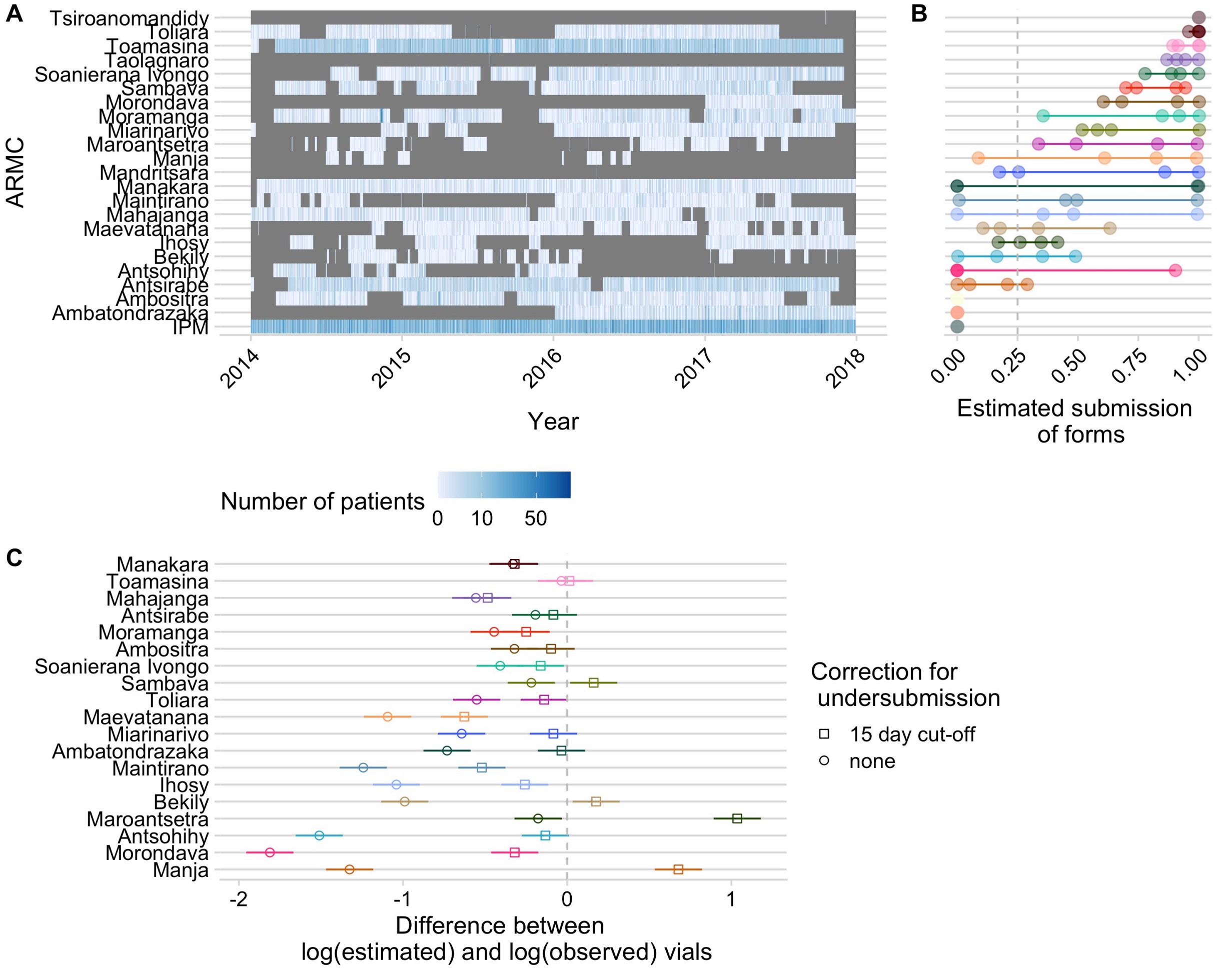

***Fig S2.2. Estimating under-submission of patient forms.***

*(A) Number of patient forms submitted to IPM for each clinic over the study period for each clinic, with periods of time where no forms were submitted for >= 15 days excluded (in grey); (B) Estimates for the proportion of forms submitted for each clinic (points are the estimate for each year and the line is the range), calculated as the # of days in a year which were not excluded based on the criteria of 15 consecutive days of non-submission/365. (C) The difference between log(estimated) and log(observed) vials provisioned for the period of 2013 - 2017 for each clinic correcting for under-submission (squares) using the 15 day cut-off show in A and B, vs. not correcting for under-submission (circles). We did not have data on vials provisioned for IPM.*

***Table S2.1. Root mean squared error (MSE) between observed vials provisioned and estimated by the different consecutive day threshold for correcting for periods of no form submission, with the minimum root MSE in bold.***

| Consecutive day threshold | Root MSE |
|---|---|
| 1 | 2346.38 |
| 5 | 1038.45 |
| 10 | 1015.88 |
| **15** | **1006.82** |
| 30 | 1063.39 |
| No correction | 1658.48 |

### S3. Modeling reported bite incidence

We used a weakly informative prior for the intercept of the models centered around the mean of the bite incidence for the given dataset with a standard deviation of 10. For the covariate terms, we centered the priors around zero with a standard deviation of 10. For the variance terms ($\sigma_0$ and $\sigma_e$), we set uniform priors ($unif(0,5)$). We calculated the Deviance Information Criterion (DIC, a metric of model fit to data) for each candidate model as well as the maximum potential scale reduction factors (psrf) for each covariate and the multivariate psrf for the whole model (both are metrics of model convergence, where values < 1.1 are indicative of convergence, Table S3.1) [2].

To test how well our models predicted the data, we sampled parameter estimates from the posterior distribution for each model to generate predictions to compare to data. In addition, we used the models to predict out-of-fit data (i.e. estimates from models fitted to the national data were used to predict the Moramanga data, and estimates from models fitted to the Moramanga data were used to predict the national data). Finally, to check how correcting for incomplete submission of forms affected our modeling results, we fitted our final models to the raw data uncorrected for submission (i.e. assuming forms were completely reported resulting in lower estimates of bite incidence) and with a lower cut-off (7 days, resulting in higher estimates of under-submission of forms).

For the national data, including a catchment random effect improved predictions (Fig S3.2 & Fig S3.3). However, after accounting for overdispersion, catchment effects were not clearly identifiable (Table S3.1) and the models resulted in similar predictions (Fig S3.6 & S3.7), indicating that catchment effects could not be differentiated from random variation in the data. Similarly, while the commune model fit to the Moramanga data generated stronger travel time effects (Fig 4B), after accounting for data overdispersion, the posterior estimates of the parameters overlapped for the commune and district models fit to the national data (Fig S3.4), and the model estimates were in general less robust to overdispersion than for the national data, particularly at low travel times (Fig S3.5). Population size alone was the poorest fit to the data as estimated by DIC (Table S3.1), and models with population size as an additional covariate did not generate realistic predictions to the observed data or when used to predict out of fit (Figs S3.2 and S3.3).

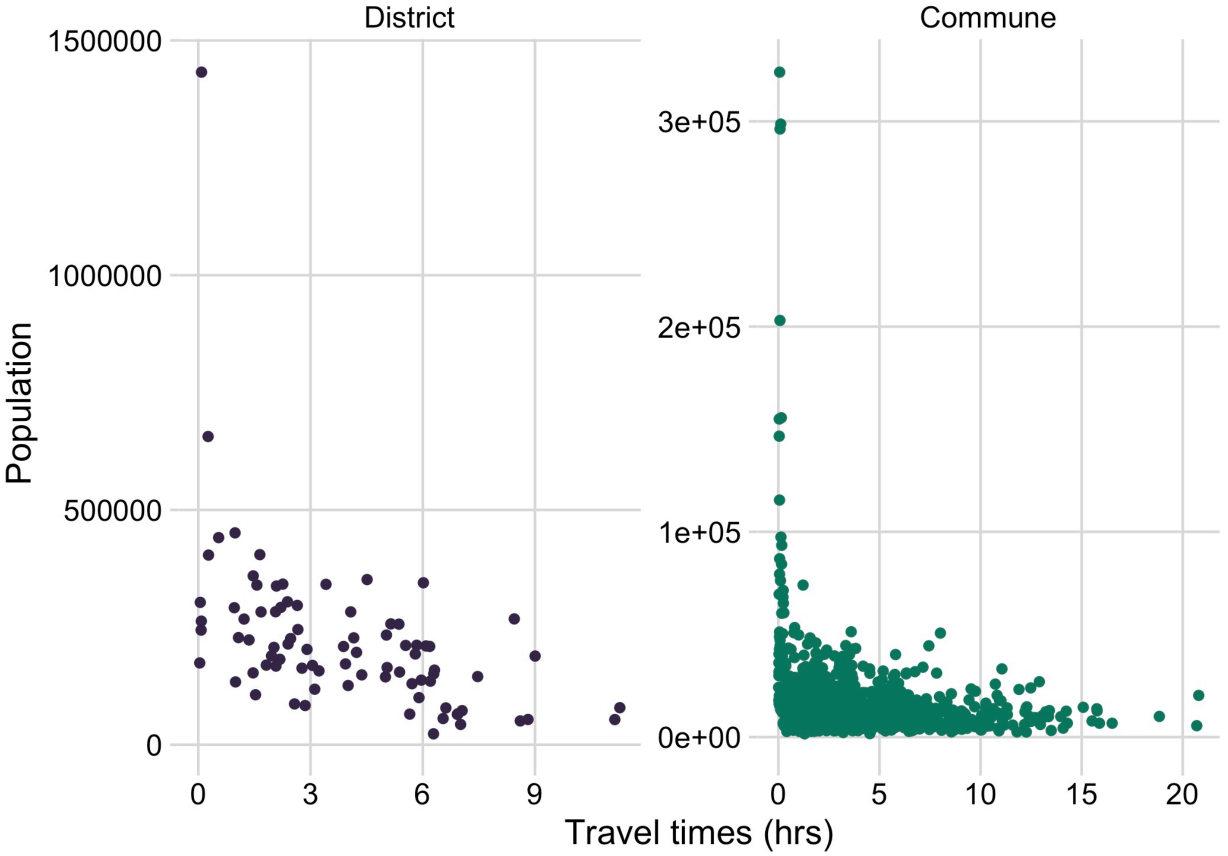

***Fig S3.1. Correlation between travel time in hours (the average weighted by the population) and population size of administrative units at the district and commune scale.***

***Table S3.1. DIC and convergence estimates (maximum potential scale reduction factor and multivariate psrf, values < 1.1 indicate convergence) for all models.*** *For the column pop effect, addPop = models with population size as additional covariate, onlyPop = models with population as only covariate, flatPop = models with population as offset in model. For the intercept type: random = random intercept by catchment, fixed = a single fixed intercept was estimated). The Overdispersion column indicates whether an overdispersion parameter was estimated (yes) or not (no).*

| Dataset | Scale | Pop effect | Intercept type | Overdispersion | DIC | Max psrf | Multivariate psrf |
|---|---|---|---|---|---|---|---|
| Moramanga | Commune | flatPop | fixed | no | 10.664 | 1.001 | 1 |
| Moramanga | Commune | onlyPop | fixed | no | 12.122 | 1.001 | 1 |
| **Moramanga** | **Commune** | **flatPop** | **fixed** | **yes** | **2.721** | **1.062** | **1.017** |
| Moramanga | Commune | addPop | fixed | no | 8.425 | 1.011 | 1.003 |
| National | Commune | addPop | fixed | no | 119.917 | 1.002 | 1.001 |
| National | Commune | onlyPop | random | no | 144.383 | 1.001 | 1.001 |
| National | Commune | onlyPop | fixed | no | 213.415 | 1 | 1 |
| National | Commune | flatPop | random | no | 41.936 | 1.001 | 1.001 |
| National | Commune | addPop | random | no | 50.287 | 1.001 | 1.001 |
| **National** | **Commune** | **flatPop** | **fixed** | **yes** | **6.784** | **1.028** | **1.008** |
| National | Commune | flatPop | random | yes | 6.793 | 1.39 | 1.102 |

| | | | | | | | |
|---|---|---|---|---|---|---|---|
| | | | | | | 3 | |
| National | Commune | flatPop | fixed | no | 89.813 | 1.001 | 1 |
| National | District | flatPop | fixed | no | 113.715 | 1.001 | 1 |
| National | District | addPop | fixed | no | 124.957 | 1.001 | 1 |
| National | District | onlyPop | random | no | 126.176 | 1.002 | 1.001 |
| National | District | onlyPop | fixed | no | 189.105 | 1.001 | 1 |
| National | District | flatPop | random | no | 59.12 | 1.001 | 1.001 |
| **National** | **District** | **flatPop** | **fixed** | **yes** | **6.781** | **1.324** | **1.087** |
| National | District | flatPop | random | yes | 6.783 | 1.122 | 1.069 |
| National | District | addPop | random | no | 61.133 | 1.004 | 1.001 |

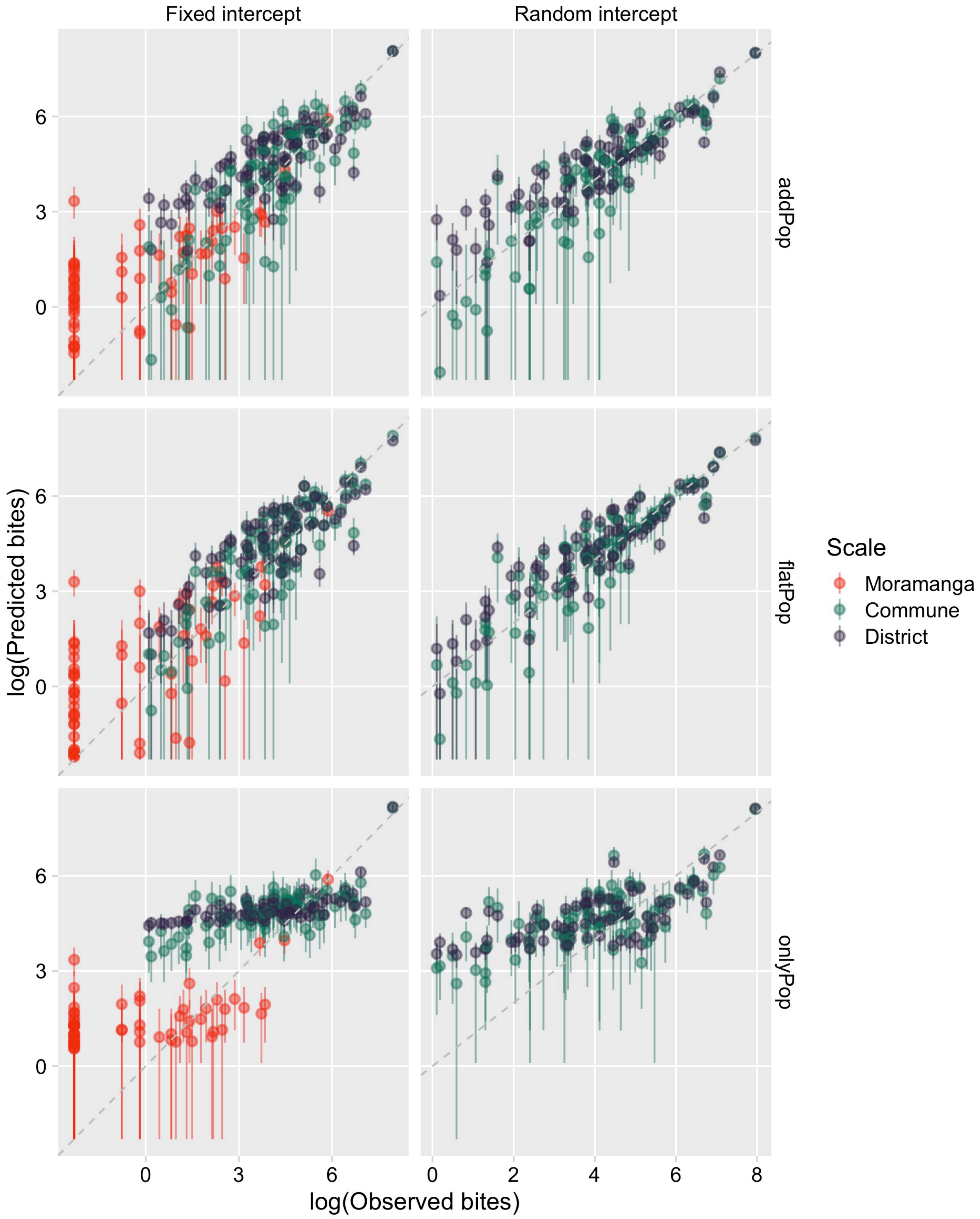
Fixed intercept
Random intercept
addPop
flatPop
onlyPop
log(Predicted bites)
log(Observed bites)
Scale
Moramanga
Commune
District
0
3
6
0
2
4
6
8

***Fig S3.2. Prediction to data used to fit each model.***

*Log of the observed bites against the log of predicted bites generated from sampling 1000 independent draws from the posterior distributions for each parameter, with the points the mean of the predictions and the linerange the 95% prediction intervals. Columns are by the type of model intercept (either a fixed intercept or a random intercept by catchment) and rows are the type of model structure with respect to the population covariate (addPop = population size as additional covariate, onlyPop = population as only covariate, flatPop = population as offset in model). Colors show which data set was used for fitting and the scale of the model (Moramanga = Moramanga data with covariates at the commune level, Commune = National data with covariates at the commune level, District = National data with covariates at the district level).*

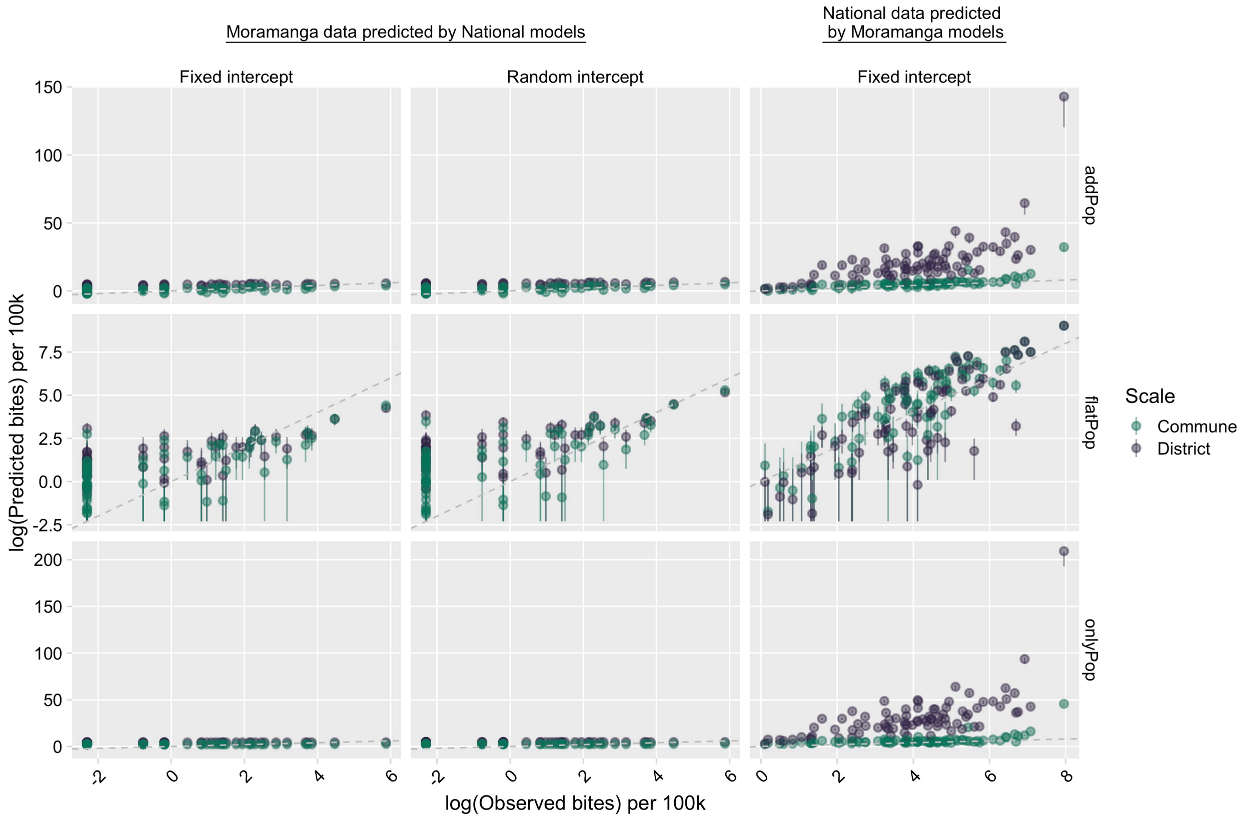

***Fig S3.3. Out of fit predictions to data.***

*Log of the observed bites against the log of predicted bites for data not used to fit the model. Predictions were generated by sampling 1000 independent draws from the posterior distributions for each parameter, with the points the mean of the predictions and the linerange the 95% prediction intervals. The first two columns are the predictions from the commune and district model fitted to the national data for the Moramanga data with fixed and random intercepts. The third column are predictions from models fitted to the Moramanga data for the national data at the commune and district scale (only fixed intercept models). Rows are the type of model structure with respect to the population covariate and colors show which data set was used for fitting as per Fig S3.2.*

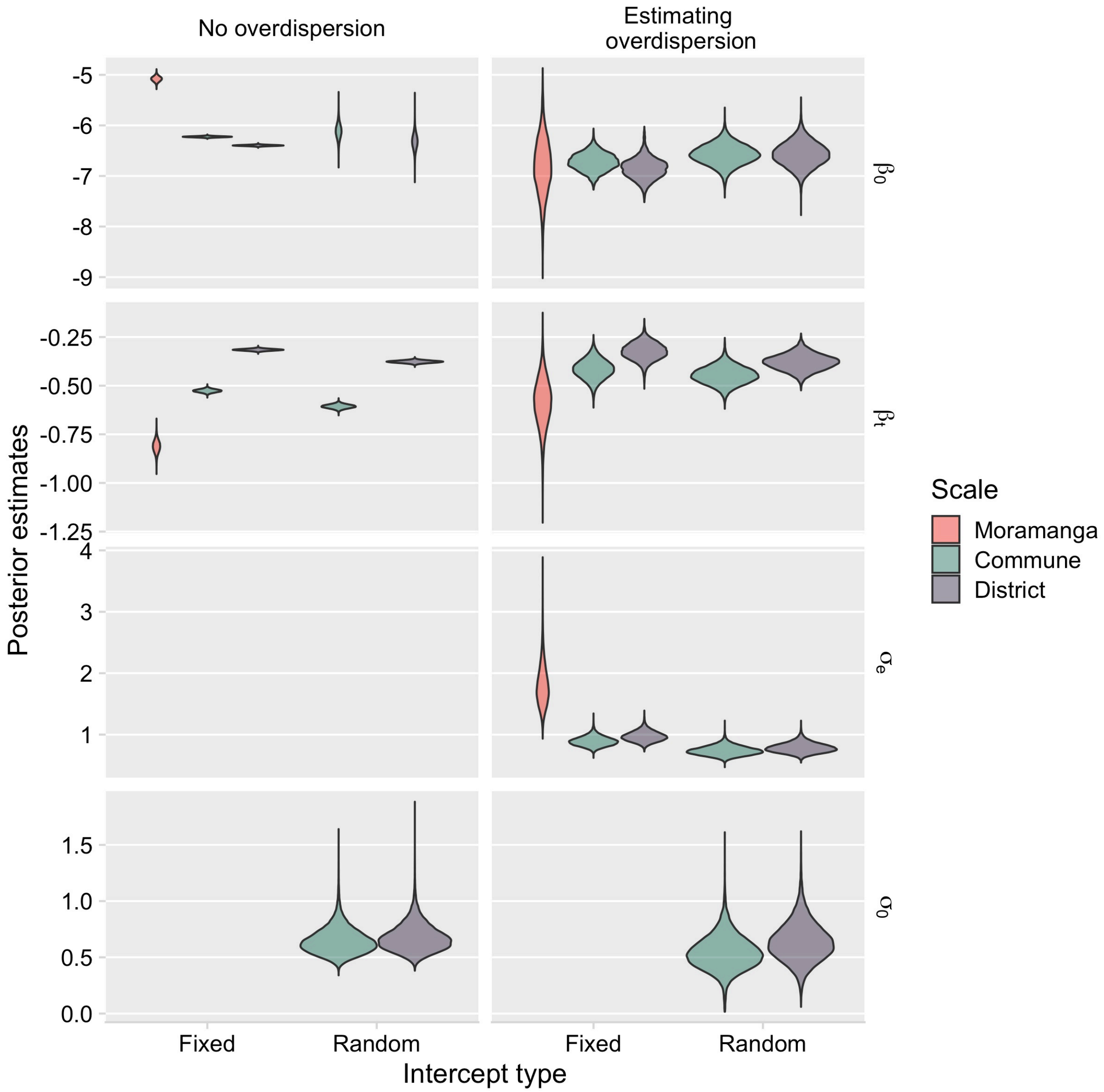

***Fig S3.4. Posterior estimates of parameters from models with travel time and population as an offset.***

*Comparing models accounting for overdispersion ($\sigma_e$) compared to models with no overdispersion parameter (flatPop in Figs S3.2 & S3.3). For the Moramanga model, as data came from a single catchment, models with a random catchment effect ($\sigma_0$) were not fitted.*

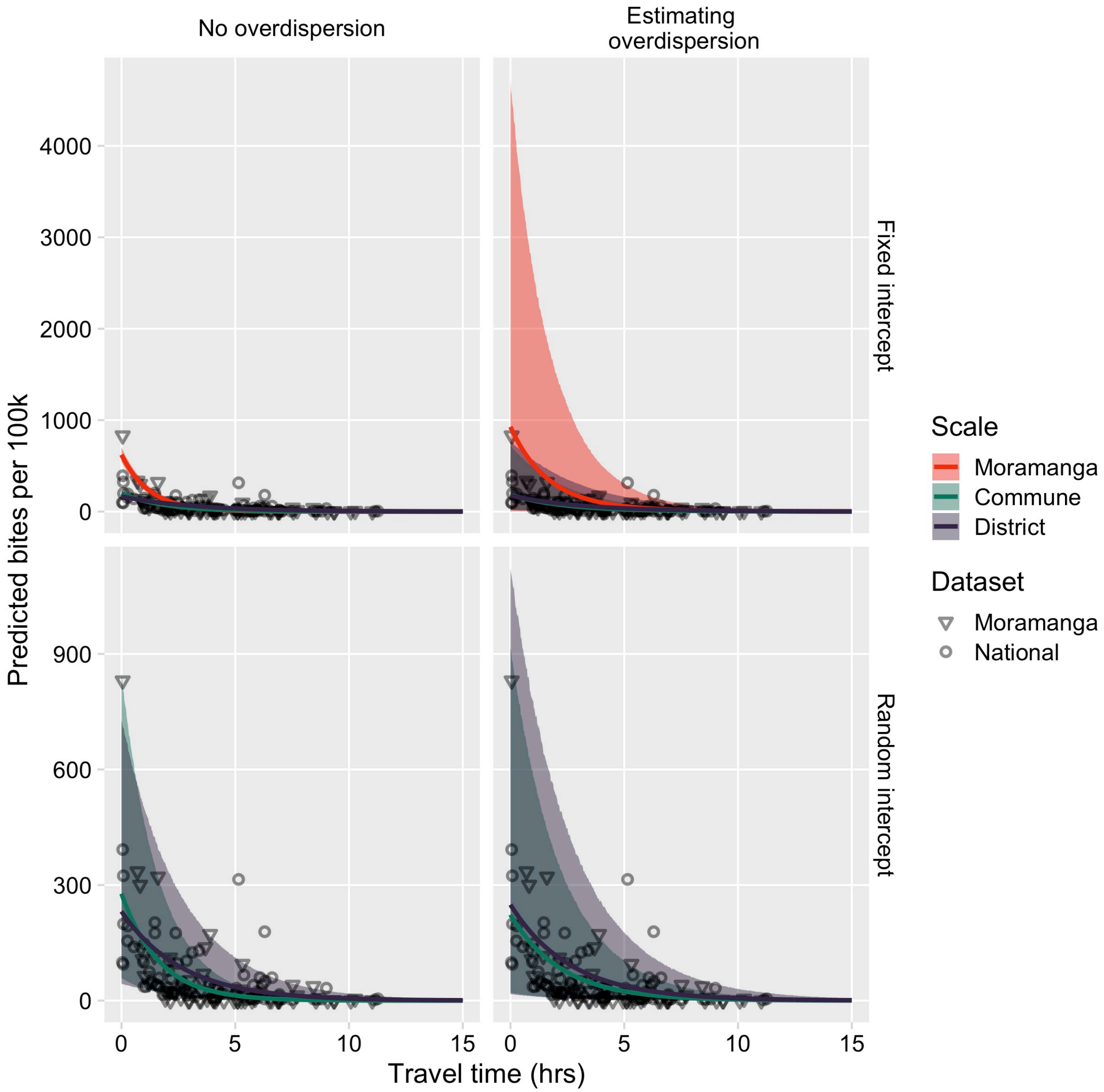

***Fig S3.5. Predicted relationship between travel times (in hours) and reported bite incidence per 100,000 persons.***

*Generated from sampling 1000 independent draws from the posterior distributions for each parameter, with the line the mean of the predictions and the envelopes showing the 95% prediction intervals. Rows are by the type of model intercept (either a fixed intercept or a random intercept by catchment) and columns are whether the model estimated an overdispersion parameter. The points show the data used to fit the models (the National dataset), as well as the Moramanga dataset. Note the different y-axis limits between the fixed and random intercept models.*

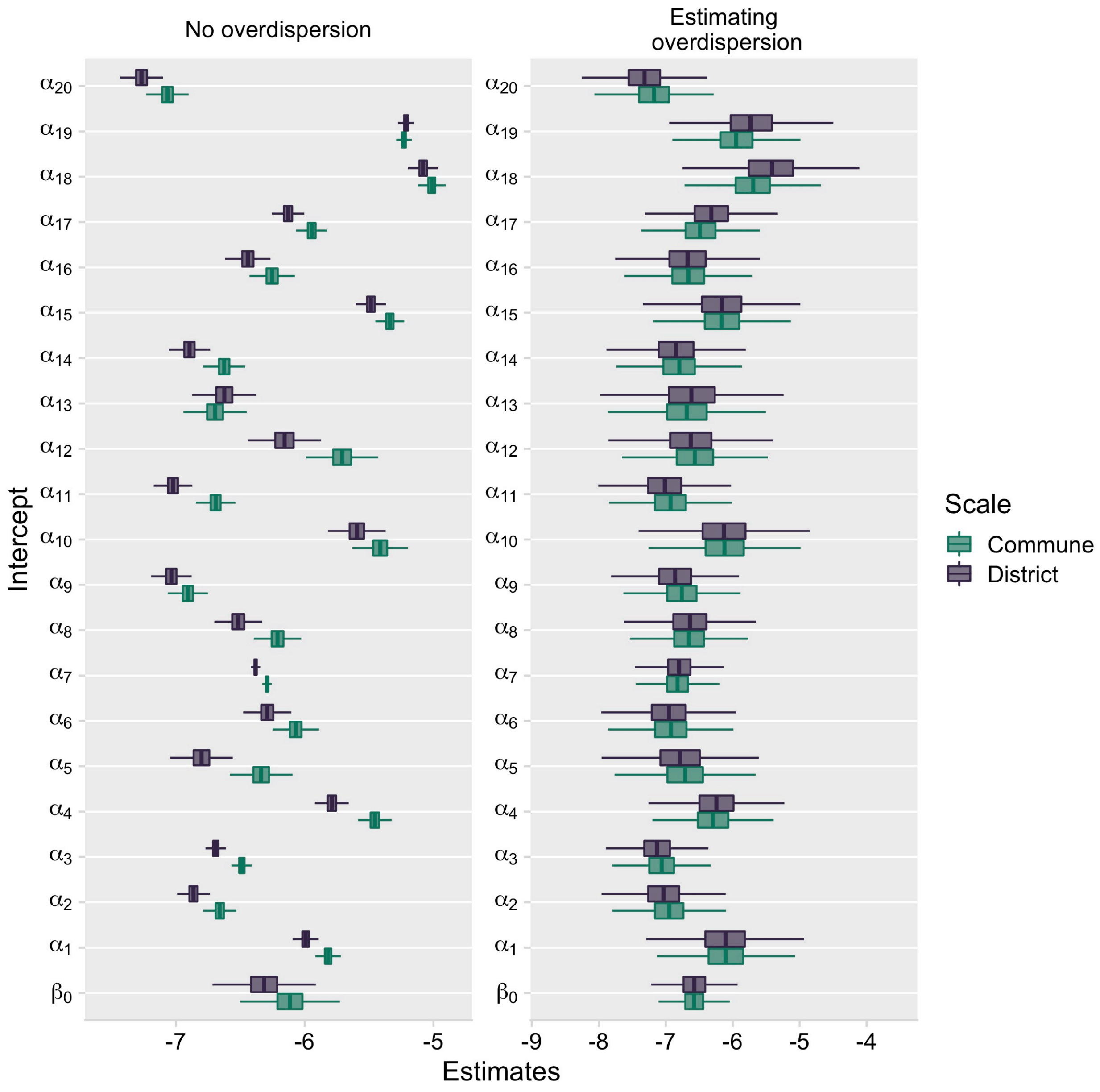

***Fig S3.6. Posterior estimates of the catchment intercepts ($\alpha$ parameters, with $\beta_0$ the estimated mean intercept) for models with and without an overdispersion parameter.***

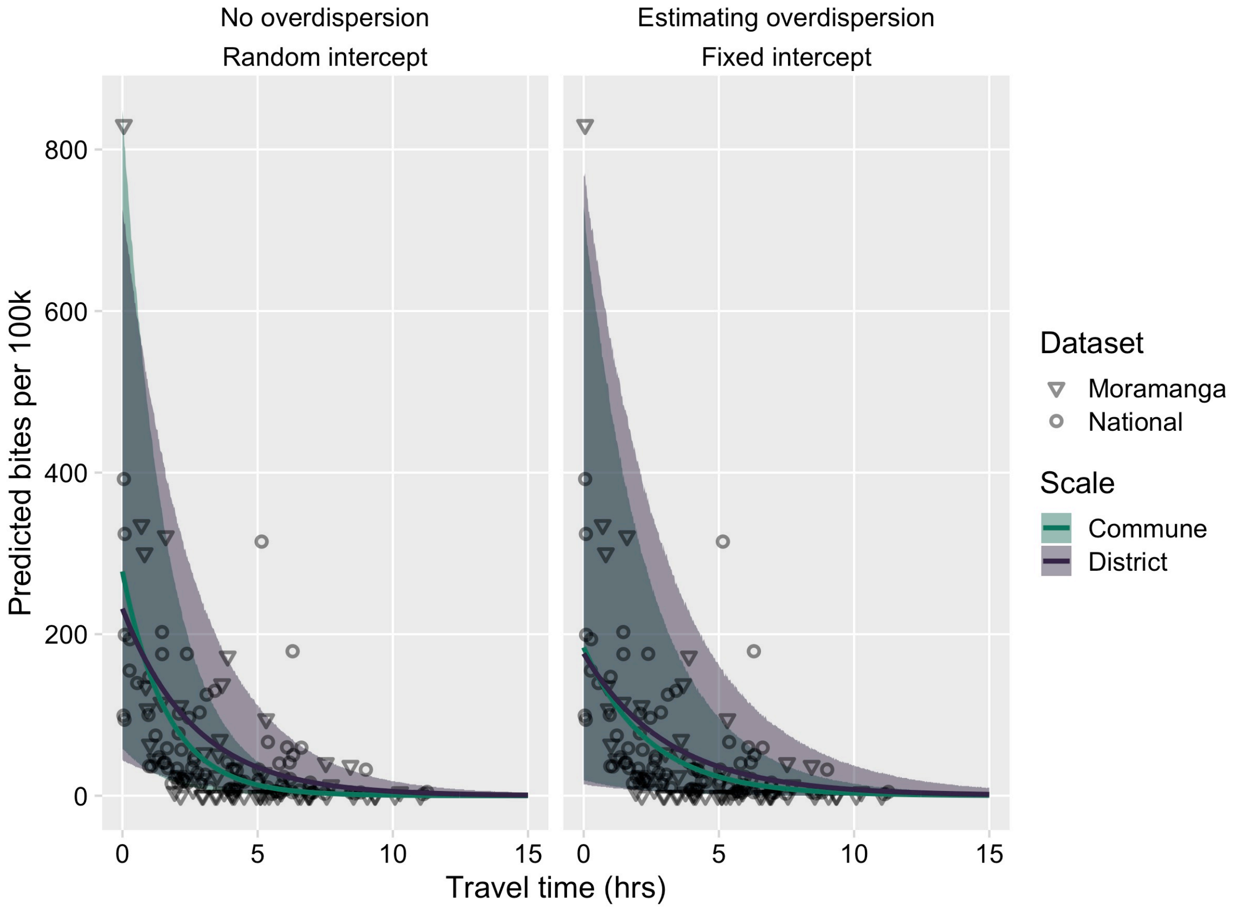

***Fig S3.7. Predicted relationship between travel times (in hours) and reported bite incidence per 100,000 persons.***

*For random intercept model without overdispersion vs. fixed intercept model with overdispersion, generated from sampling 1000 independent draws from the posterior distributions for each parameter, with the line the mean of the predictions and the envelopes showing the 95% prediction intervals. The points show the data used to fit the models (the National dataset), as well as the Moramanga dataset.*

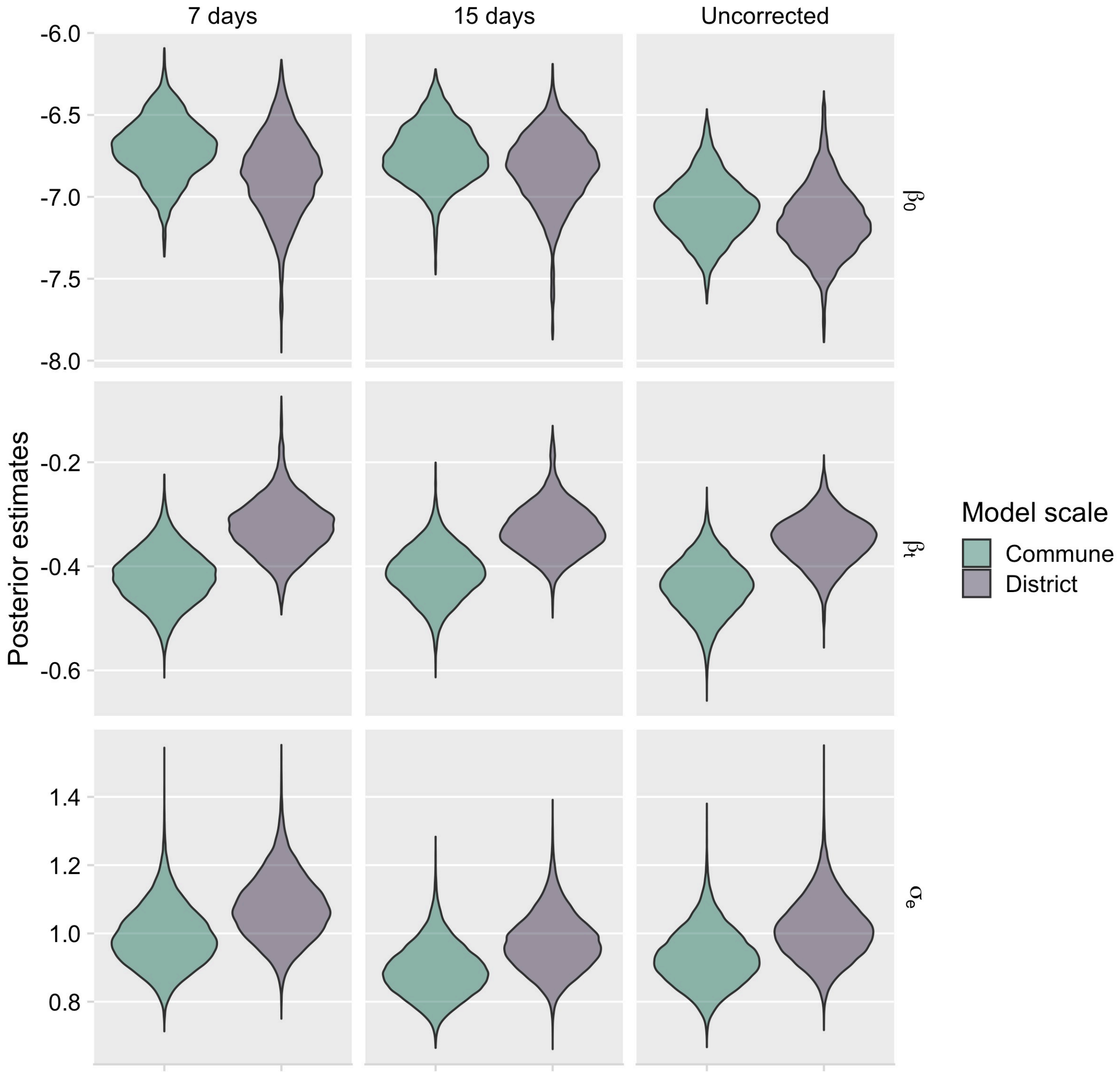

***Fig S3.8. Posterior estimates for models with population as an offset and an overdispersion parameter.***

*Columns show estimates from models fitted to the national dataset (1) corrected for both under-submission by correcting for periods of at least 7 days where zero patient forms were submitted, (2) correcting for periods of 15 days where zero patient forms were submitted, and (3) with the raw data not correcting for under-submission.*

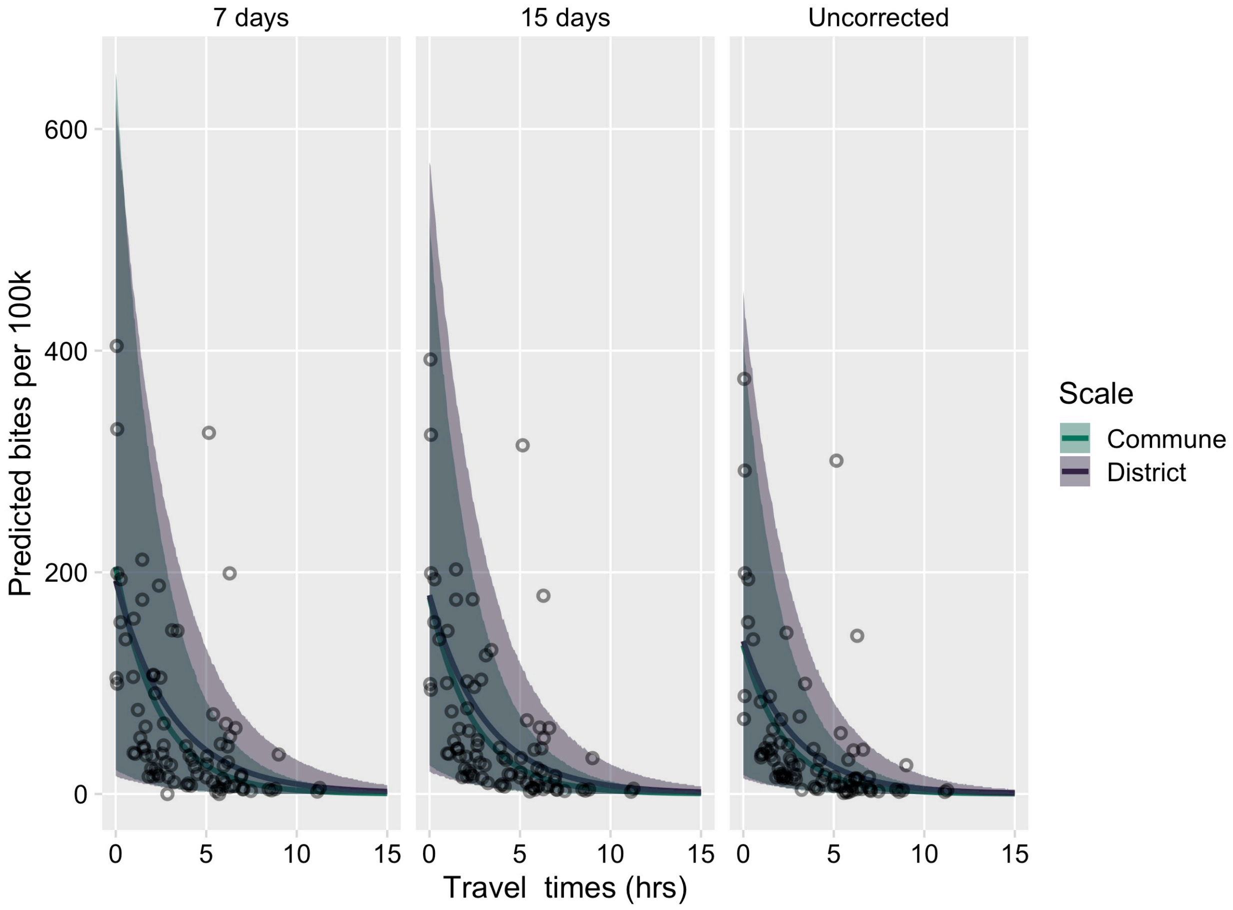

***Fig S3.9. The estimated relationship between travel time in hours (x-axis) and mean annual bite incidence per 100,000 persons (y-axis).***

*For models with population as an offset and an overdispersion parameter. Panels show predictions from models fitted to the national dataset 1) correcting for periods of at least 7 days where zero patient forms were submitted (a less stringent cutoff resulting in lower estimates of the proportion of forms submitted and thus higher estimates of reported bite incidence), (2) correcting for periods of 15 days where zero patient forms where submitted, as presented in the main analysis, and (3) with the raw data not correcting for under-submission (resulting in lower estimates of reported bite incidence). Predictions were generated from sampling 1000 independent draws from the posterior distributions for each parameter, with the line the mean of the predictions and the envelopes showing the 95% prediction intervals. The points show the data used to fit the models.*

### S4. Range of rabies exposure incidence in people

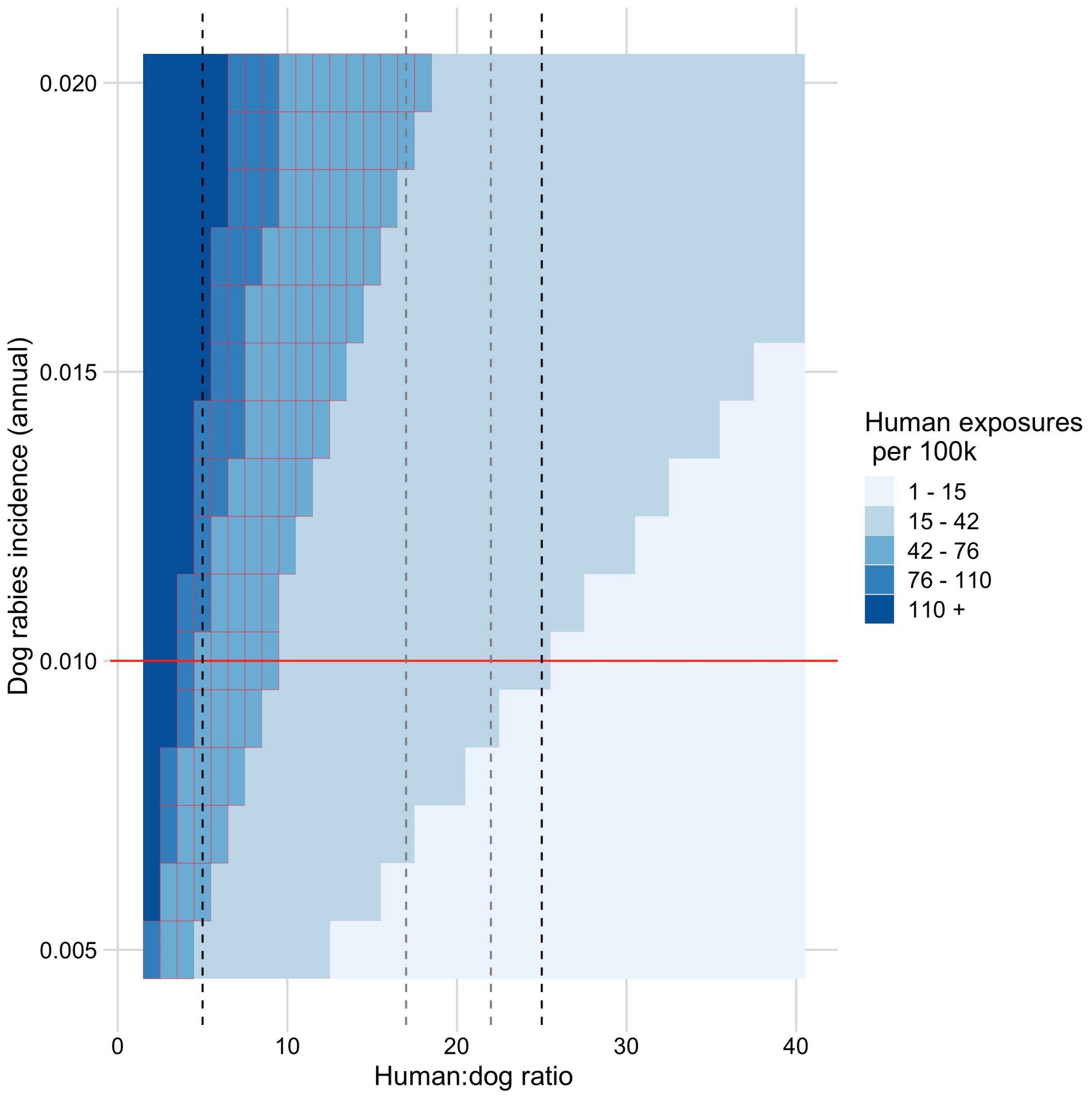

***Fig S4.1. Estimated exposures per 100,000 given a range of human to dog ratios (HDRs, x-axis) and annual dog rabies incidence (y axis).***

*Assuming that each dog on average exposes 0.39 persons [3]. The black dashed lines show the range of human to dog ratios (HDRs) we use in the main analysis to estimate the range of human exposure incidence (where the red horizontal line and black dashed lines intersect). The grey dashed lines show the HDRs estimated from the Moramanga district from a recent study [4]. The cells with red outlines show the range of estimated exposure incidence from a previous study of bite patients in Moramanga District [1].*

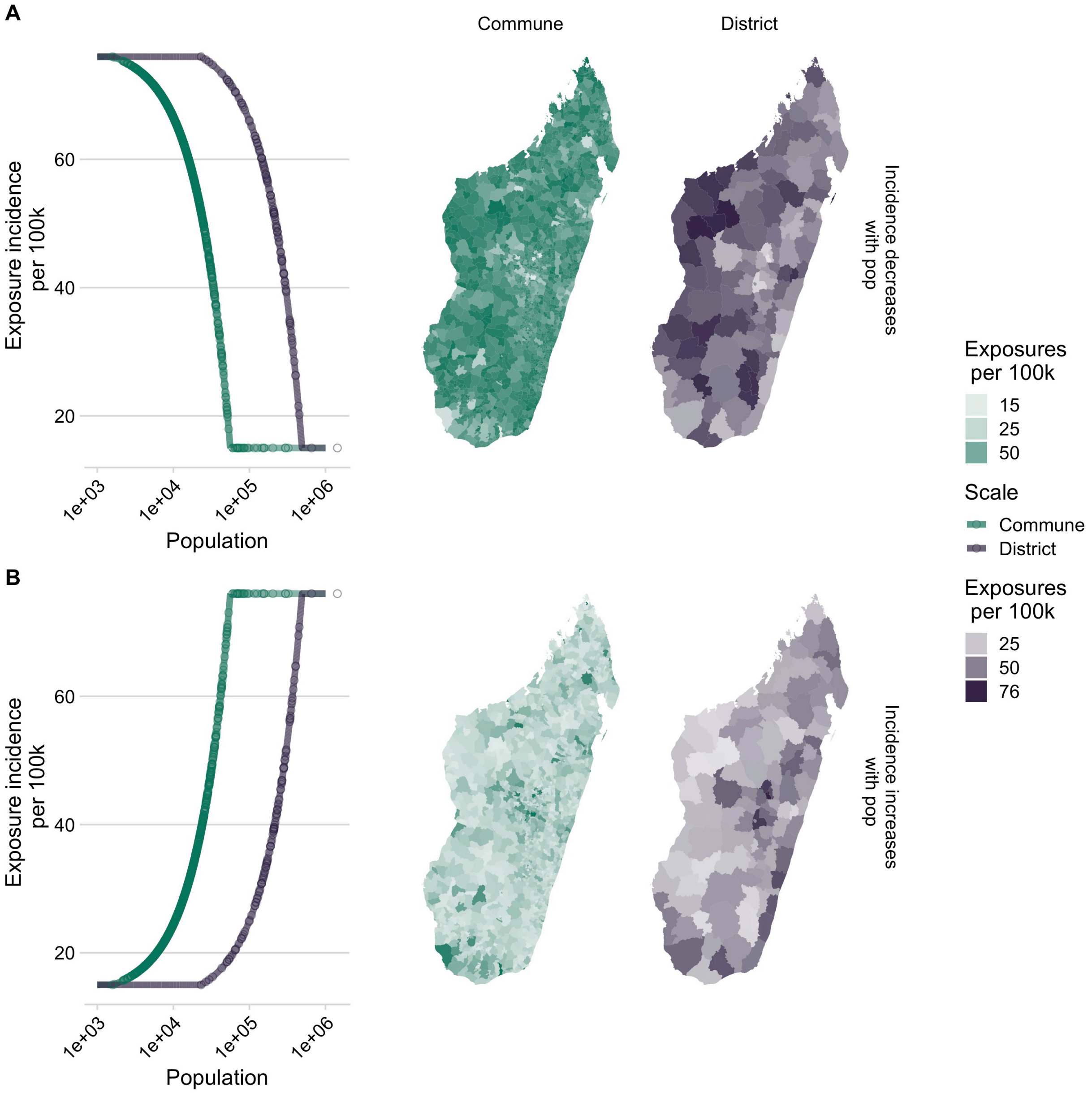

***Fig S4.2. Range of constrained scaling factors generated for district and commune population size***

*Underlying rabies exposures either (A) decreases with increasing population size or (B) increases with increasing population size across a fixed range of exposure incidence (15.6 - 76 exposures/100k persons). Lines show the expected relationship, with points showing where administrative units fall along this curve, and maps show how this results in variation in assumed exposure incidence spatially at the commune and district level.*

**S5. Estimating the impact of expanding PEP provisioning to additional clinics**

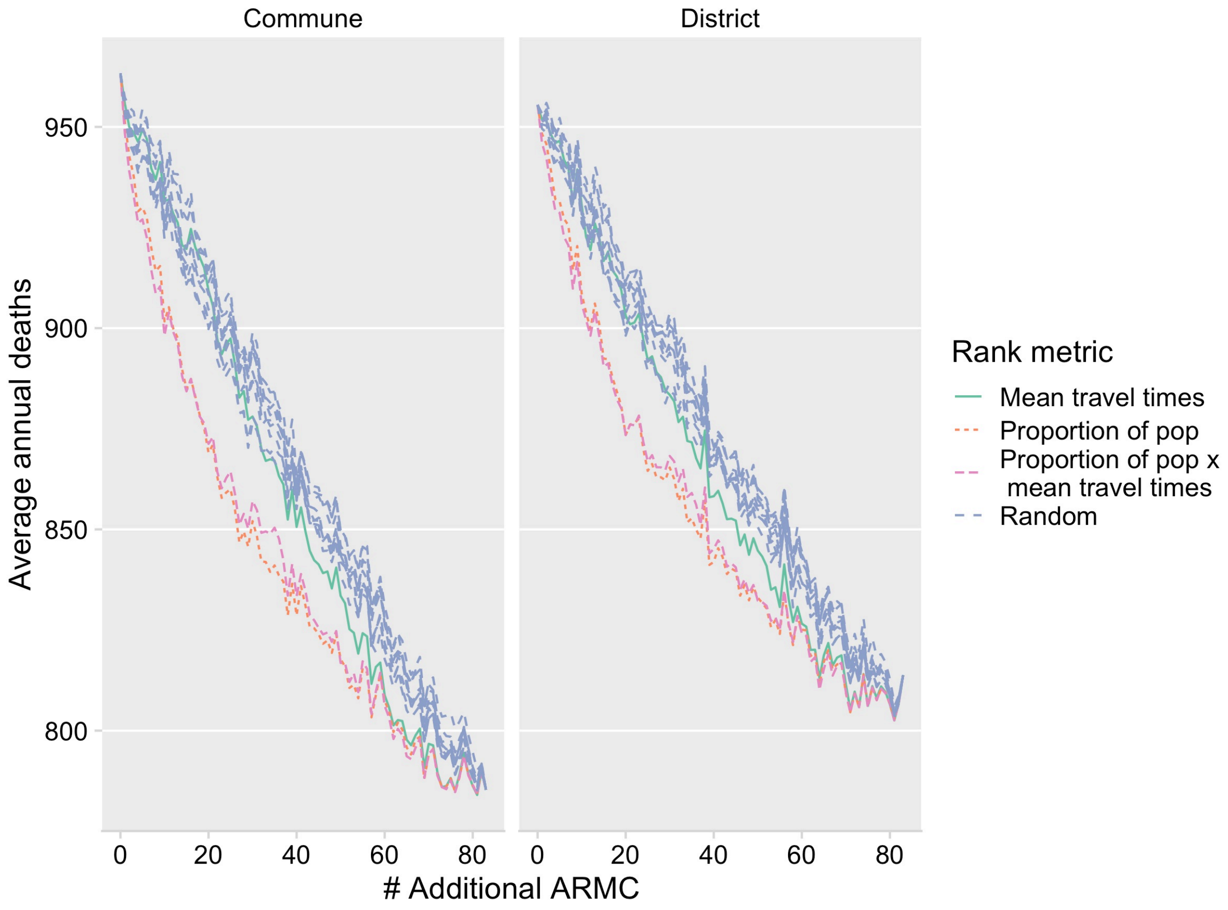

***Fig S5.1. Comparing metrics for ranking clinics for targeted expansion.***
*We simulated expansion using three different ranking metrics: 1) reduction in mean travel times (green line) 2) the proportion of the population for which travel times were reduced (red dashed line) and 3) the proportion of the population for which travel times were reduced weighted by the reduction in travel times (pink dashed line). For each of these, we simulated burden using our decision tree framework (y axis is the mean of 1000 simulations of annual deaths at the national level). The blue lines show 10 simulations of randomly expanding access on reducing burden as a comparator. The panels show to the commune and district model of reported bite incidence.*

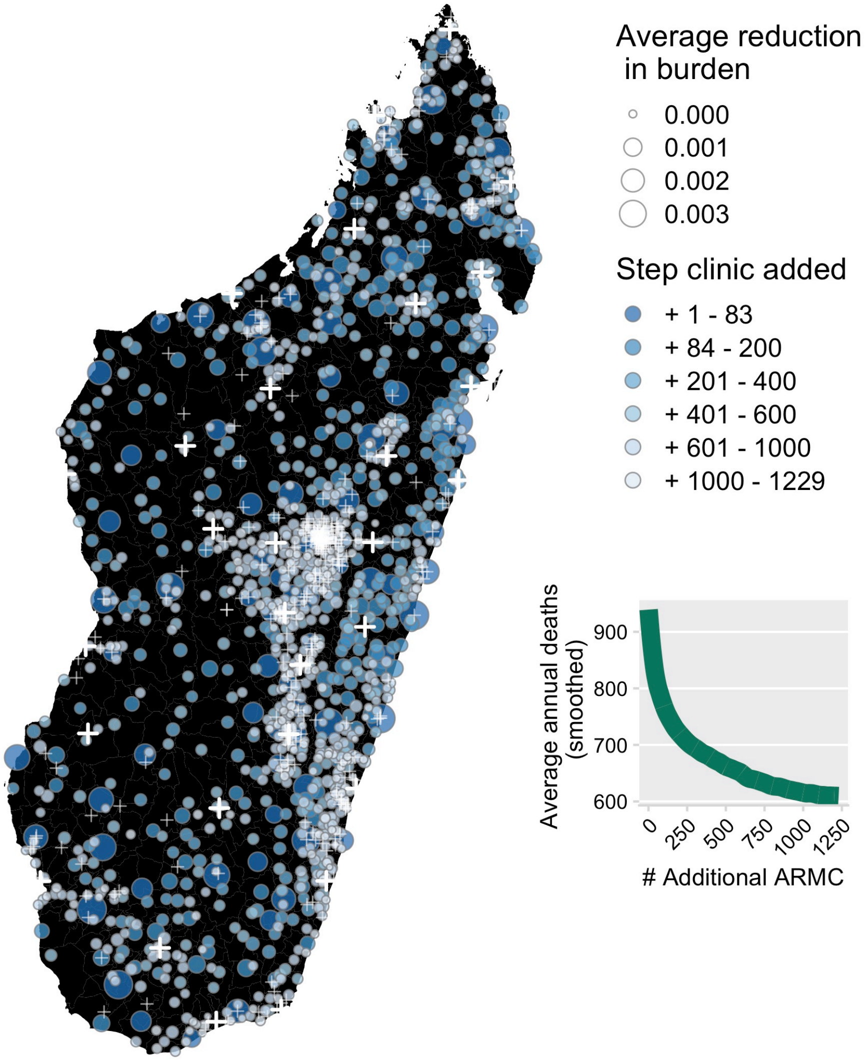

***Fig S5.2. Map of the location and at what step each clinic was added.***

*The circles are each of the primary clinics across the country sized by the resulting average reduction in burden (based on smoothed annual burden estimates from the commune model, see inset). The large white crosses show the location of the existing 31 clinics provisioning PEP in Madagascar and the smaller white crosses are the additional primary clinics in the country which were added in the final step but not ranked.*

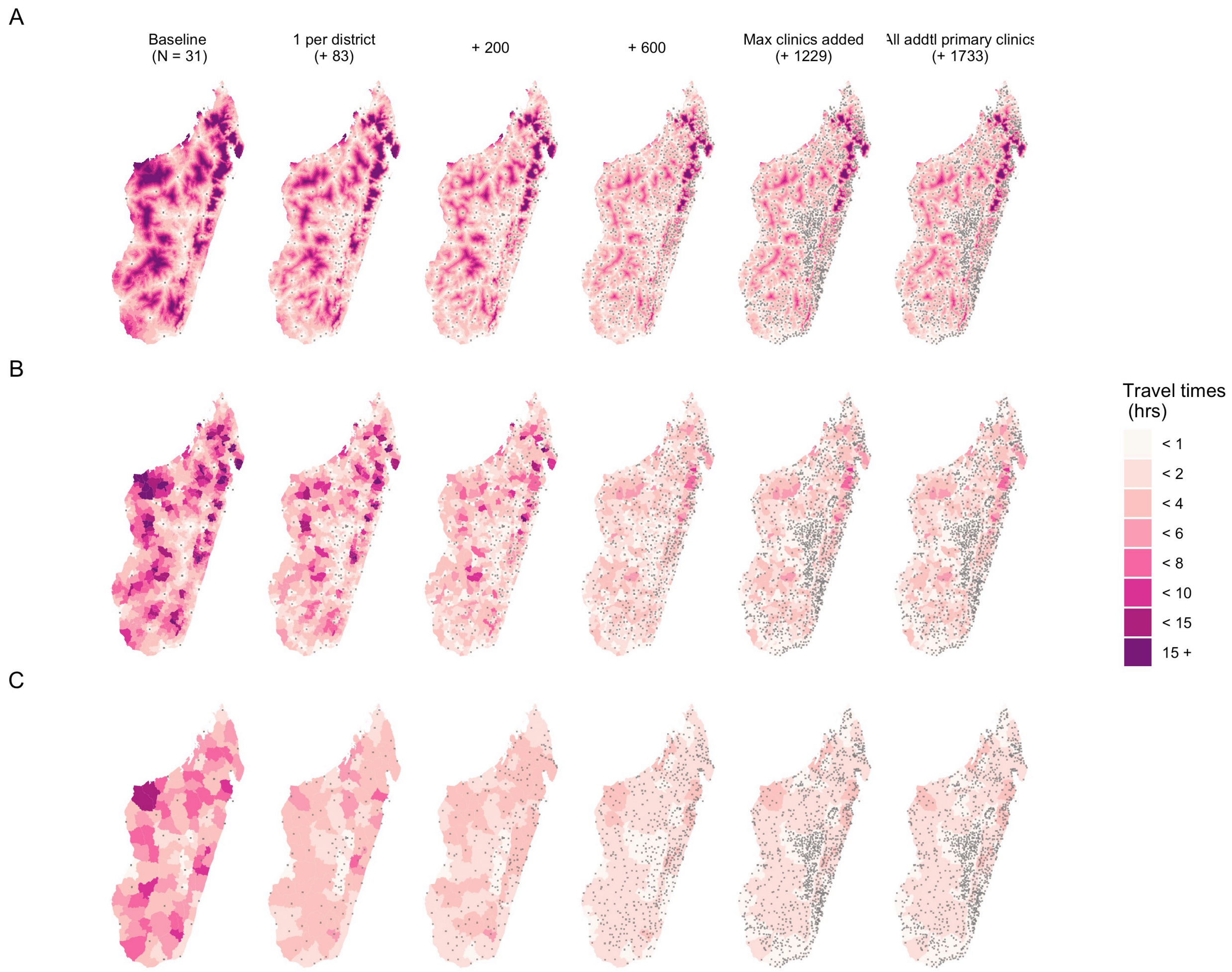

***Fig S5.3. Maps of how travel times change as clinics are added.***

*(A) at the ~1 x 1 km grid cell (B) commune and (C) district scales. The columns are ordered by the number or clinics at each step: baseline (N = 31), + 83 (1 per district), + 200, + 600, + 1406 (1 per commune), and max (+ 1696 clinics, all additional primary clinics in the country). Grey pixels show the location of clinics provisioning PEP at each step. Commune and district values are the average grid cell travel times weighted by the population in each cell.*

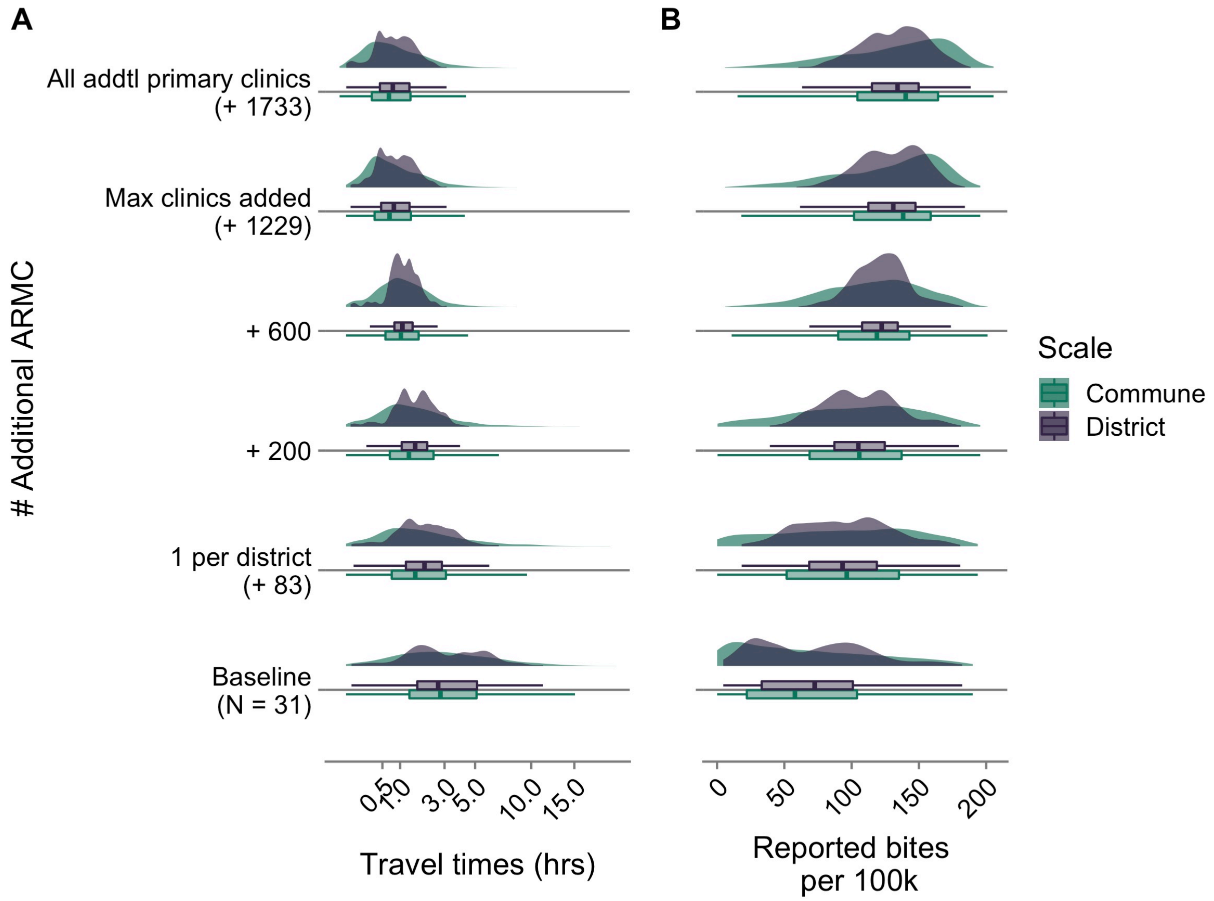

***Fig S5.4. Shifts in key metrics as clinics are added.***

*(A) travel times (hrs, x-axis is square root transformed), (B) bite incidence per 100,000 persons and boxplots showing the median for communes and district models (colors).*

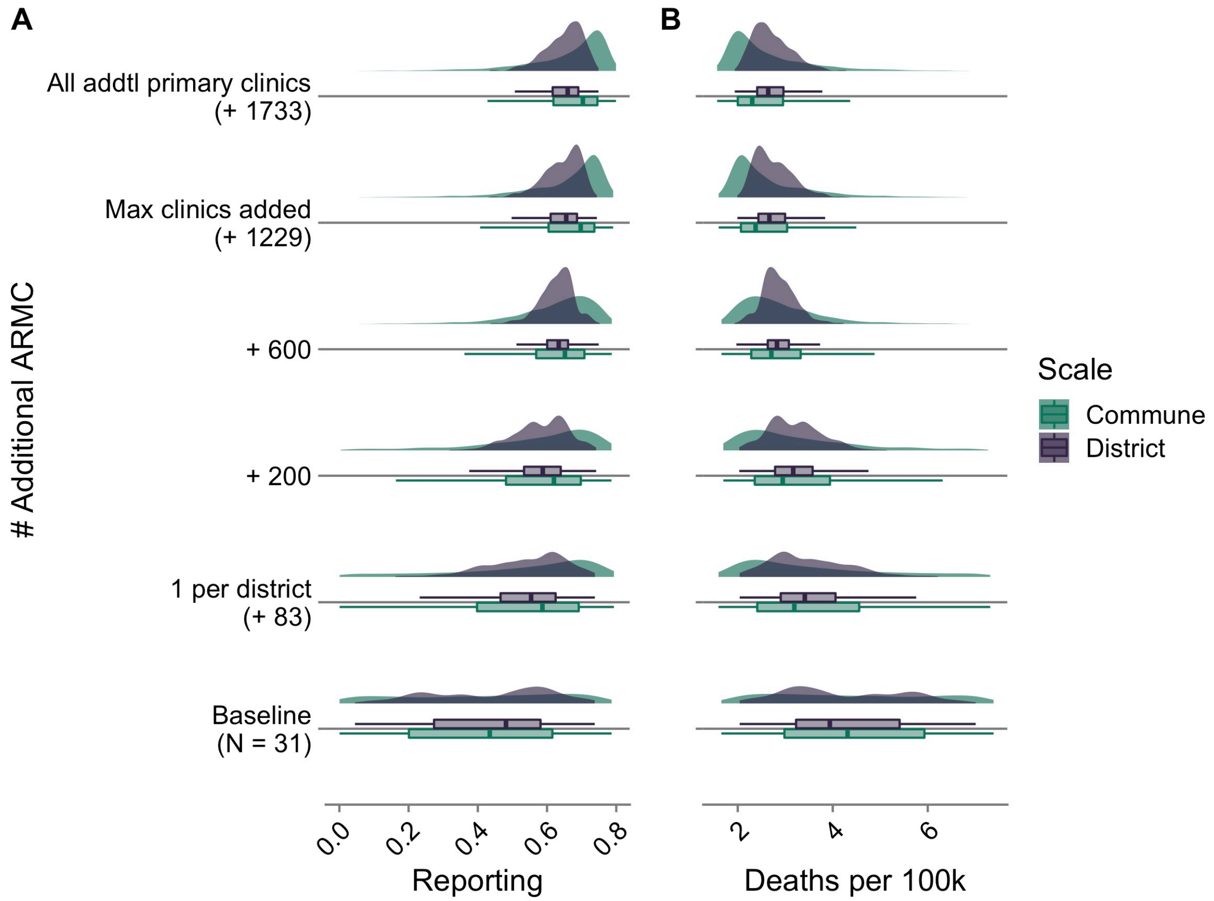

***Fig S5.5. Shifts in key metrics as clinics are added.***

*(A) reporting, (B) death incidence per 100,000 persons for the commune and district models (colors) as clinics are added.*

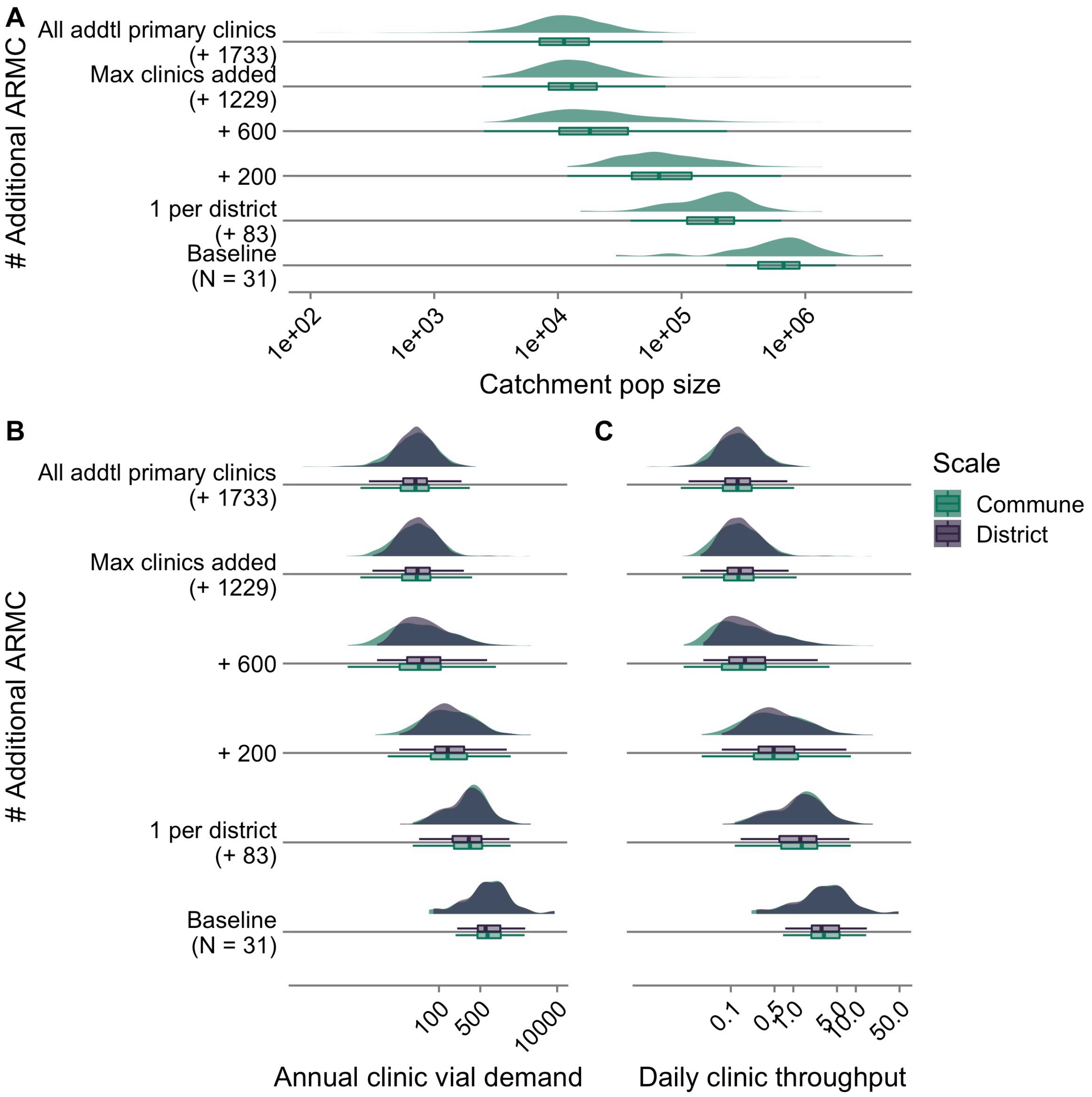

***Fig S5.6. Shifts in key metrics as clinics are added.***

*(A) catchment population size, (B) annual vial demand, and (C) daily throughput (i.e. average number of patients reporting each day) given estimates of bite incidence for the commune and district models (colors). For vial demand estimation, catchment population sizes are the same for each model as these populations are allocated at the grid cell level (i.e. population in a grid cell is allocated to the clinic catchment it is closest to in terms of travel times regardless of district or commune). All x-axes are log transformed.*

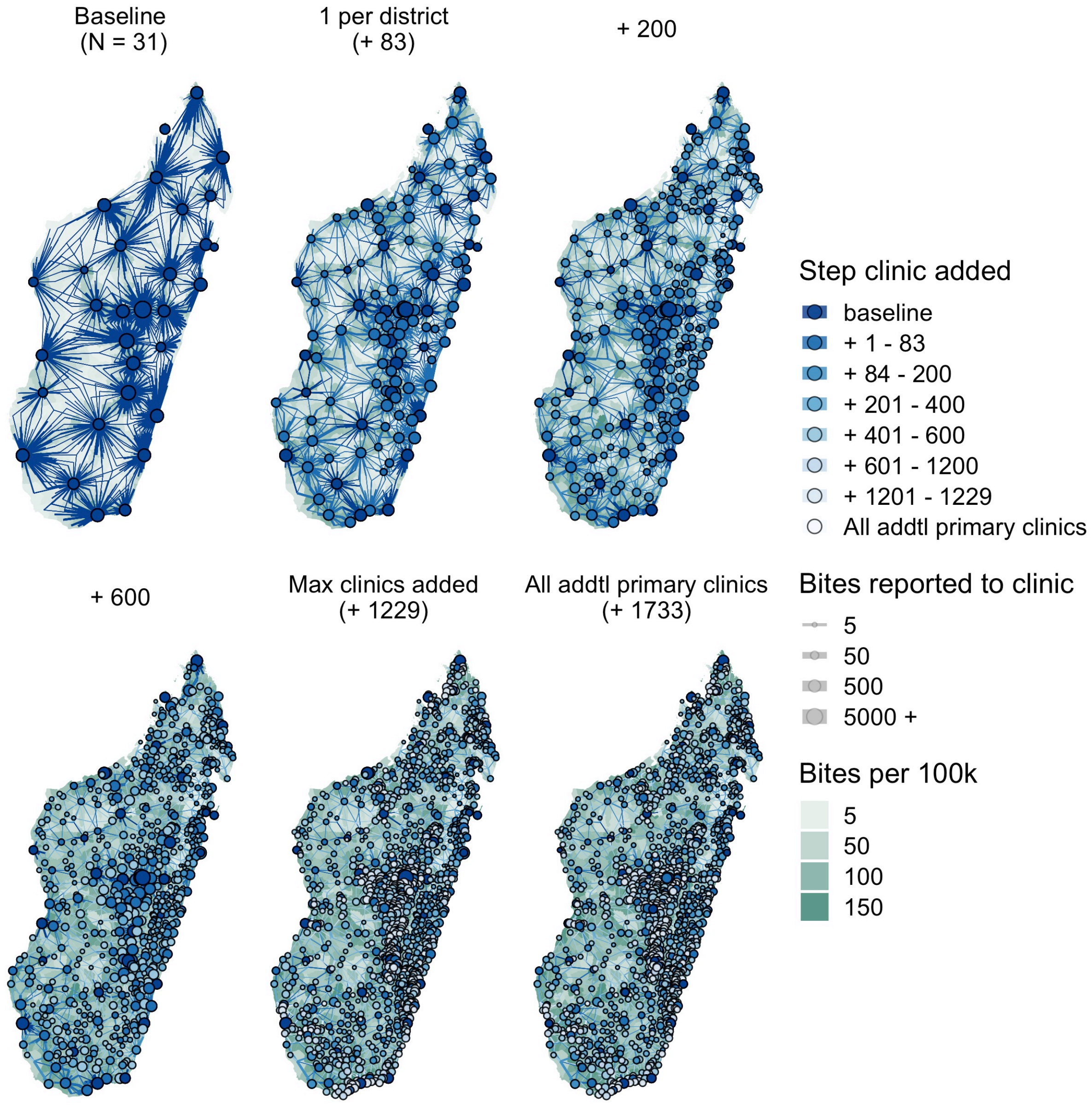

***Fig S5.7. Shifts in where bites are reported to as clinics are added for the commune model.***

*The circles show the clinic locations for each scenario, with size proportional to the annual average bites reported to that clinic. Lines show where the bites are reported from (commune centroid) also proportional to the number of bites. The polygon shading shows the commune level reported bite incidence.*

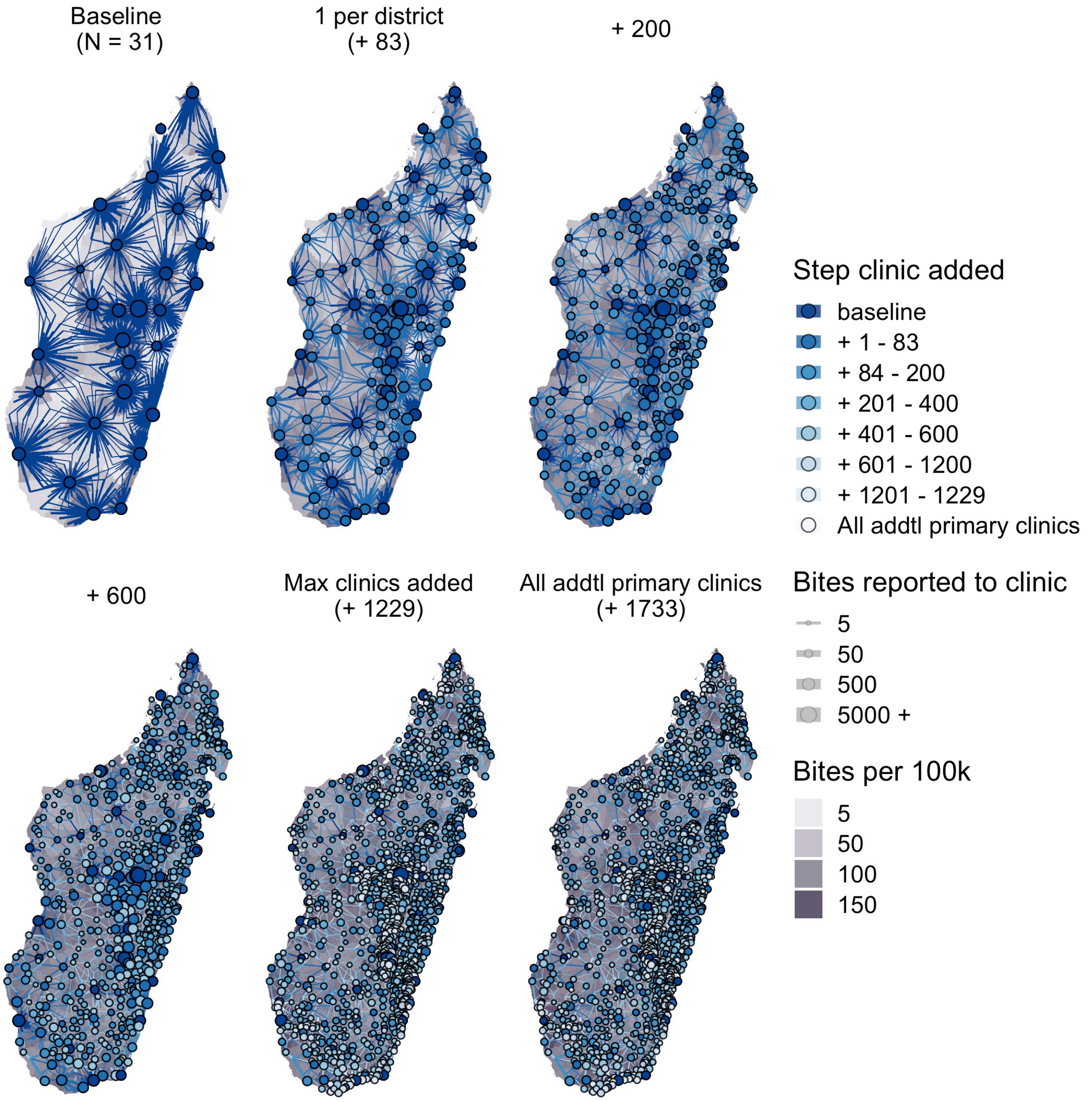

***Fig S5.8. Shifts in where bites are reported to as clinics are added for the district model.***

*The circles show the clinic locations for each scenario, with size proportional to the annual average bites reported to that clinic. Lines show where the bites are reported from (commune centroid) also proportional to the number of bites. The polygon shading shows the district level reported bite incidence.*

### S6. Sensitivity of burden estimates to parameter assumptions

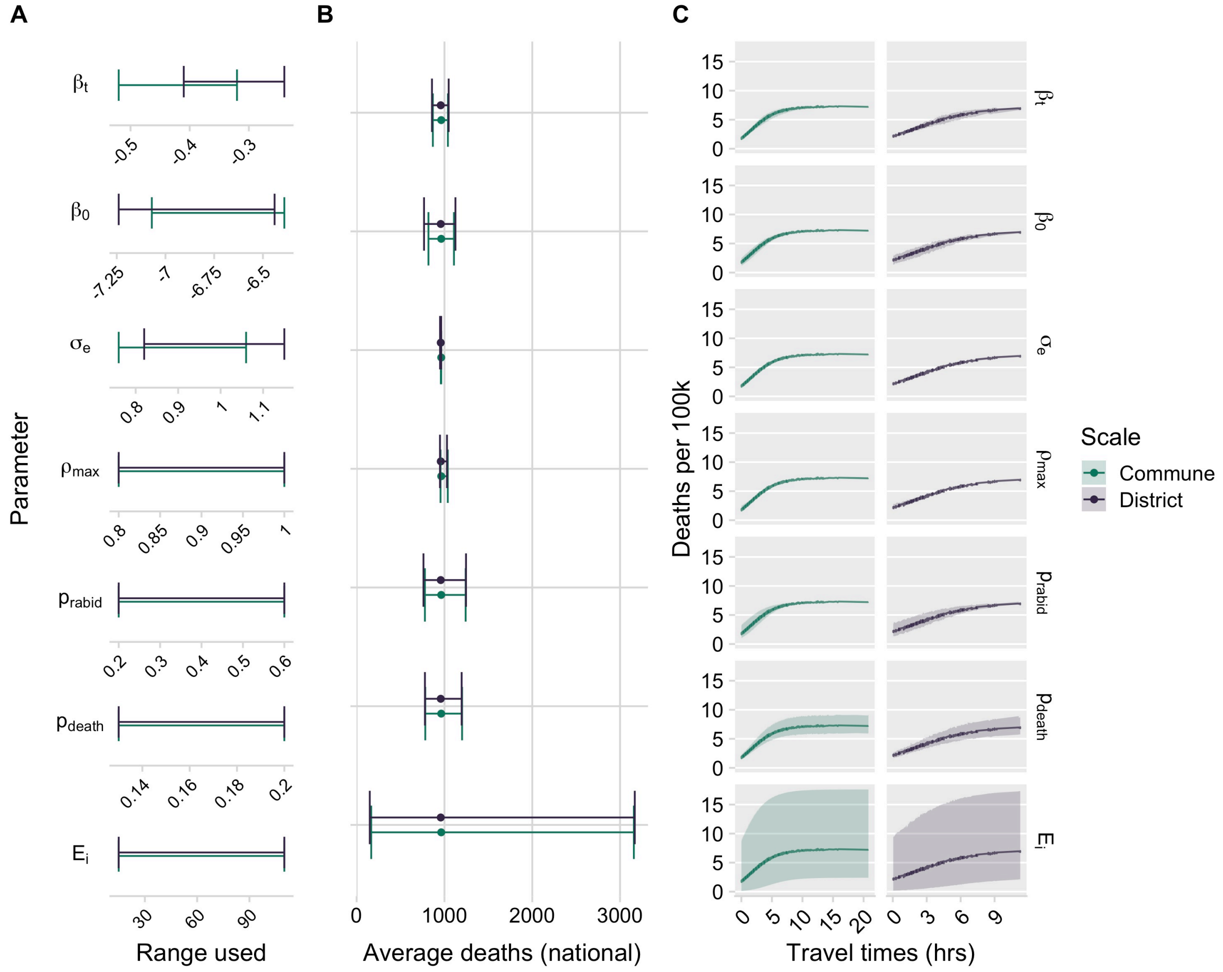

***Fig S6.1. Sensitivity analyses for baseline burden estimates.***

*(A) Range of parameters used in the univariate sensitivity analysis (upper 97.5% and lower 2.5% credible interval of posterior for the parameters in the bite incidence model, 95% of CI from 95% CI of estimate from [5] for* $\rho_m ax$*, and fixed at the minimum and maximum of the range used in the main analysis for all other parameters) (B) Range of estimates of the estimate average deaths at the national level for range of parameter estimates (point shows the estimate presented in the main analyses and the ends show the upper and lower estimates for the parameter range) and (C) estimates for the predicted relationship between travel times and incidence of human rabies deaths across the same parameter ranges (line shows the estimate presented in the main analyses and the envelope shows the upper and lower estimates for the parameter*

*range) for the two models (colors). See Table S6.1 & 2 for ranges used for each parameter.*

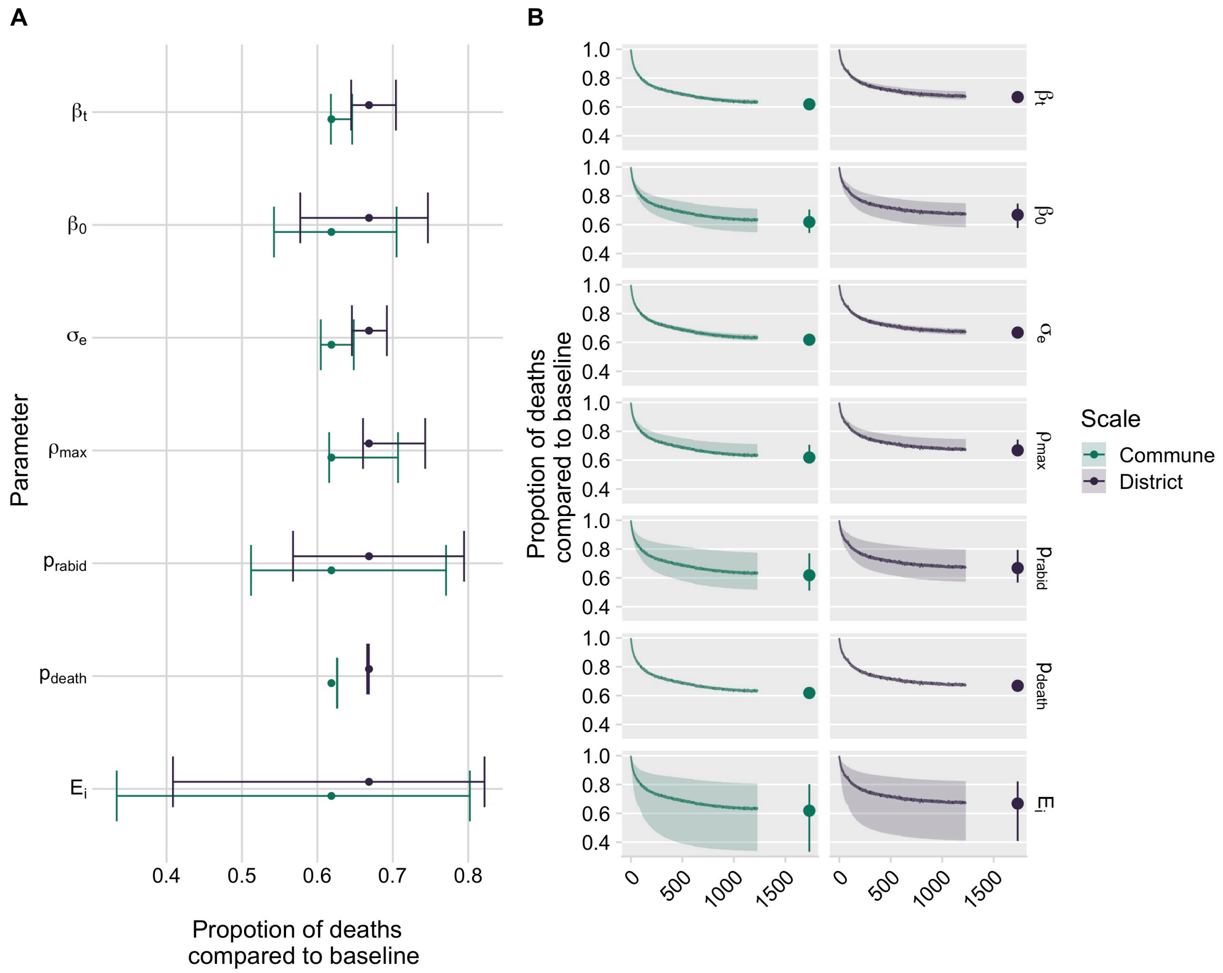

***Fig S6.2. Sensitivity analysis for PEP expansion.***

*(A) Estimates of the maximum proportion reduction of deaths from the baseline for range of parameter estimates (point shows the estimate presented in the main analyses and the ends show the upper and lower estimates for the parameter range) and (B) estimates for how human rabies decreases proportional to the baseline as clinics are added for these same parameter ranges (line shows the mean estimate presented in the main analyses and the envelope shows the mean estimates for the parameters fixed at the upper and lower range) for the two models (colors). See Fig S6.1A for ranges used for each parameter.*

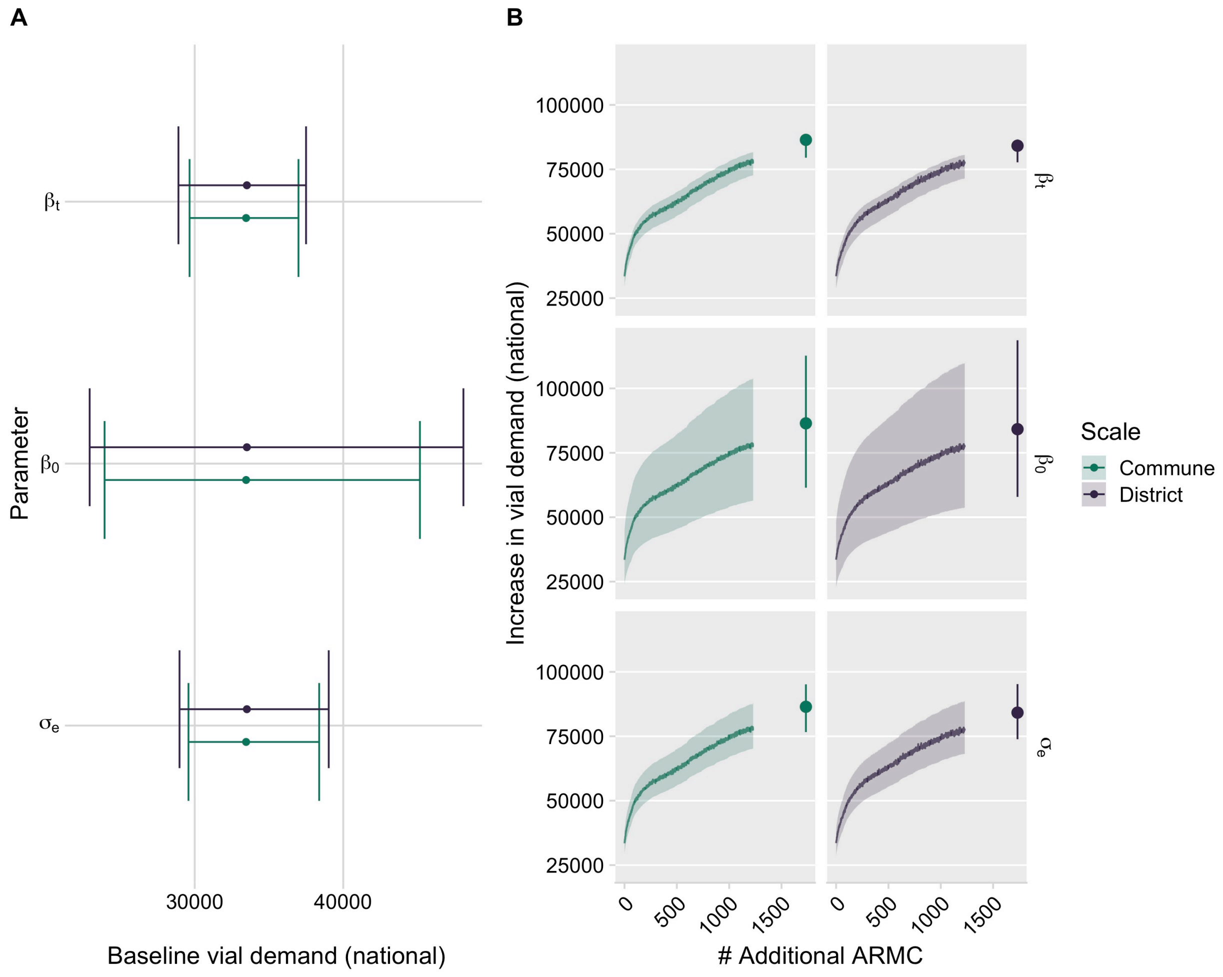

***Fig S6.3. Sensitivity analysis for vial demand.***

*(A) Estimates of the baseline vial demand for where each parameter was set to the upper and lower estimat of parameter estimates (point shows the estimate presented in the main analyses and the ends show the upper and lower estimates for the parameter range) and (B) estimates for how vial demand increases as clinics are added for these same parameter ranges (line shows the mean estimate presented in the main analyses and the envelope shows the mean estimates for the parameters fixed at the upper and lower range) for the two models (colors) for the two models (colors). See Table S6.1 & 2 for ranges used for each parameter.*

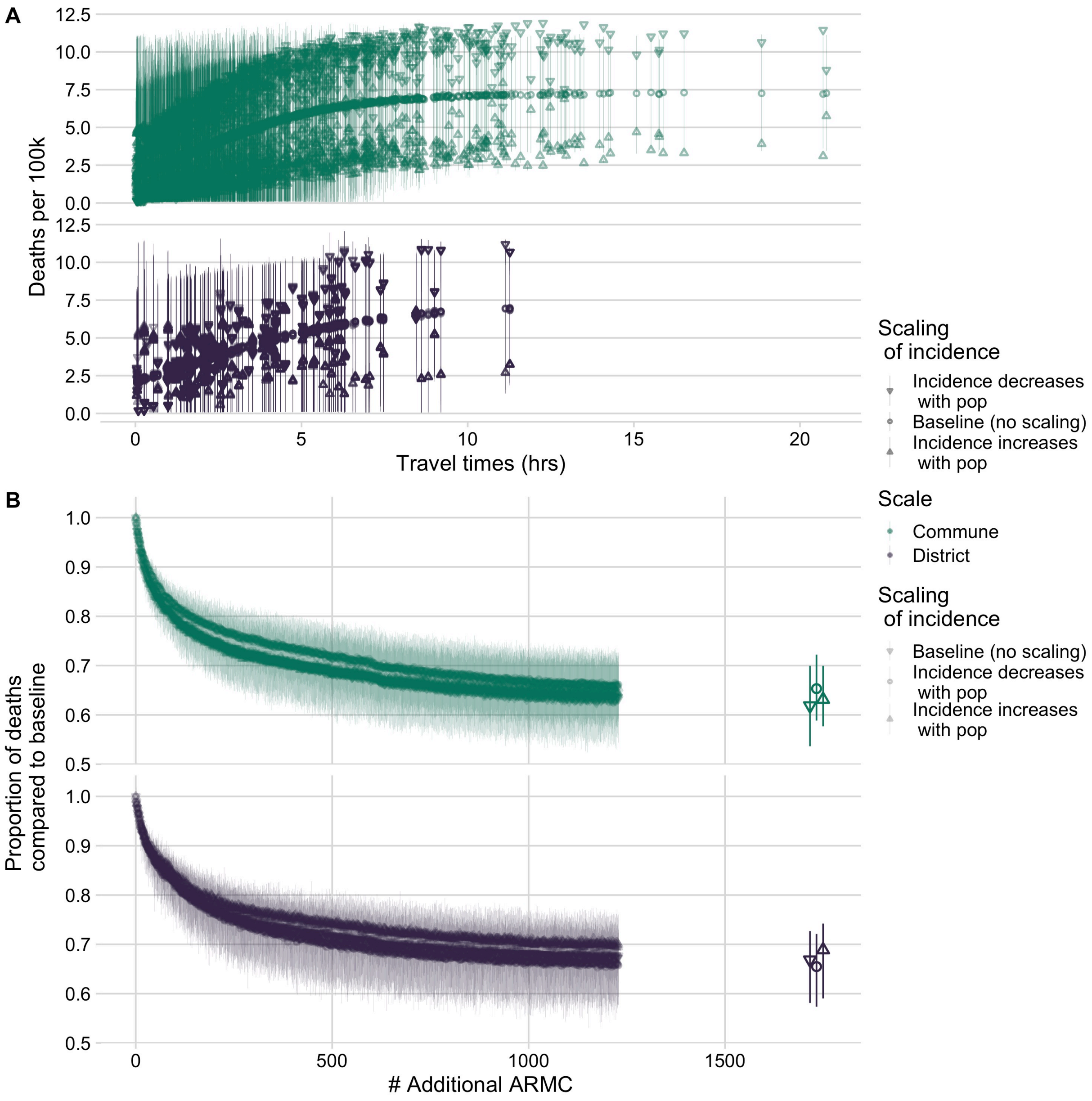

***Fig S6.4. Sensitivity analysis for exposure scaling with population.***

*Predicted relationship between (A) travel times and rabies death incidence per 100k persons and (B) the proportional reduction in deaths from the baseline as clinics are added for the district model vs. commune models (colors) with columns comparing the baseline (presented in main analyses) and assumptions of rabies exposure incidence increasing or decreasing with human population size (shapes). The points show the mean estimates from 1000 simulations and the line ranges show the 95% prediction intervals.*

**S7. Sotware citations**

9. Revolution Analytics, Weston S. Foreach: Provides foreach looping construct.
